# Identifying multi-resolution clusters of diseases in ten million patients with multimorbidity in primary care in England

**DOI:** 10.1101/2023.06.30.23292080

**Authors:** Thomas Beaney, Jonathan Clarke, David Salman, Thomas Woodcock, Azeem Majeed, Paul Aylin, Mauricio Barahona

## Abstract

Identifying clusters of co-occurring diseases can aid understanding of shared aetiology, management of co-morbidities, and the discovery of new disease associations. Here, we use data from a population of over ten million people with multimorbidity registered to primary care in England to identify disease clusters through a two-stage process. First, we extract data-driven representations of 212 diseases from patient records employing i) co-occurrence-based methods and ii) sequence-based natural language processing methods. Second, we apply multiscale graph-based clustering to identify clusters based on disease similarity at multiple resolutions, which outperforms k-means and hierarchical clustering in explaining known disease associations. We find that diseases display an almost-hierarchical structure across resolutions from closely to more loosely similar co-occurrence patterns and identify interpretable clusters corresponding to both established and novel patterns. Our method provides a tool for clustering diseases at different levels of resolution from co-occurrence patterns in high-dimensional electronic healthcare record data.

## Introduction

Multimorbidity, defined as the co-occurrence of two or more long-term conditions (LTCs) in one person, poses a significant challenge to health systems worldwide.^1, 2^ Having multimorbidity is associated with poorer quality of life,^3^ increased mortality,^4^ greater use of healthcare services and higher healthcare costs.^5, 6^ As a binary label, multimorbidity is a crude marker of medical complexity but there is growing evidence that distinct profiles or *clusters* of LTCs may be associated with differences in outcomes.^7–9^ Although some clusters of co-occurring conditions are clinically well-established, for example, a cluster of conditions representing metabolic syndrome,^10, 11^ the evolution of analysis methods for ‘big data’ opens up the use of routinely collected electronic healthcare records (EHRs) for identifying clusters of less commonly occurring conditions. The anticipated benefits of the identification of disease clusters are summarised by Whitty and Watt (2020), as an opportunity “to uncover new mechanisms for disease; to develop treatments; and to reconfigure services to better meet patients’ needs.”^12^

Over the last decade, many studies have been conducted to identify clusters of LTCs which co-occur together.^13, 14^ Among previous studies, ‘mental health’ and ‘cardio-metabolic conditions’ have consistently emerged as the two most replicable clusters.^13, 14^ However, current approaches to detect disease clusters suffer from limitations both in the use of data sources and in the approach to capture the multi-level complexity of disease associations. Firstly, most studies used a relatively small number of LTCs (median=16 and range=10-99 for the 51 studies reviewed by Busija *et al* (2019)) with coarse disease definitions (e.g., ‘Diabetes’). Secondly, most studies obtain only one clustering (with usually fewer than ten clusters), which may limit identification of associations between less common conditions.^13^ As is the case in unsupervised methods, it is unlikely that there is one single ‘true’ configuration of clusters, but rather that a sequence of clusterings, from fine resolutions with many clusters to coarse resolutions with few clusters, may reveal more nuanced associations, and serve different purposes. Indeed, multiscale graph-based clustering methods, such as Markov Multiscale Community Detection (MMCD), enable the identification of clusters at different resolutions directly from the structure of the data, without the need to pre-specify the number of clusters or impose a hierarchical structure.^15–17^

Recently, natural language processing (NLP) methods have emerged as a promising approach for handling the high-dimensional data found in EHRs.^18–20^ When trained on word sequences in natural language, these predictive models learn a vector representation for each word in a common space, referred to as a ‘word embedding’, which captures semantic and syntactic characteristics of each word using the context in which it is used in text. In an analogous fashion, such models can be applied to the coded data in EHRs, where medical codes are ‘words’ and EHRs are analogues to ‘documents’, to generate ‘disease embeddings’ that capture information from their occurrence and the sequences observed in real data.^18–22^ The disease embeddings can then be used to calculate the similarity between diseases for use in clustering. However, it remains unclear whether NLP methods incorporating additional information from sequences of diseases over time produce substantively different clusters to those obtained solely from co-occurrence-based methods, such as Multiple Correspondence Analysis (MCA), a dimensionality reduction method which has been used in several previous studies of multimorbidity clustering.^23–25^

In this study, we aim to identify clusters of diseases from observational data in an unsupervised manner, combining two recent approaches. Firstly, we generate disease representations applying two methods: one based only on co-occurrence (MCA), which is compared to newer NLP embedding methods that make use of code sequences. Secondly, we employ the multiscale graph-based clustering method of MMCD to identify disease clusters at different levels of resolution, based on the similarity of the obtained disease embeddings and compare against the k-means and hierarchical clustering algorithms commonly used in disease clustering.^13^ We apply these methods to a large and representative primary care EHR dataset of over 10 million patients in England and evaluate the resulting disease clusters to demonstrate that they provide clinically interpretable insights into disease associations.

## Results

### Description of the data

Of 15,256,726 patients aged 18 years or older registered in the Clinical Practice Research Datalink (CPRD) in England from 1^st^ January 2015 to 1^st^ January 2020, there were 10,579,232 (69.3%) with at least two of a pre-defined set of 212 LTCs (see Methods) and were thus included in the study. Characteristics of the eligible cohort are displayed in Supplementary Table 1. The median age was 52 (IQR: 36 – 68) years. There were more females than males (53.4% vs 46.6%) with a small number (263) recorded in CPRD as ‘indeterminate’ gender. The majority (73.0%) of people were recorded as being of White ethnicity, with 13.9% having no recorded data on ethnicity. There was a roughly even split between deciles of socioeconomic deprivation (measured by the Index of Multiple Deprivation), but with relatively fewer in the least deprived decile (9.1%). For each patient, we constructed two sequences for comparison: the first (“multiple”) used all diagnostic codes, and the second (“unique”) included a code only at its first occurrence. Using the unique code sequences, the median number of codes per patient was 5 (IQR: 3 – 9); using multiple code sequences, the median was 13 (IQR: 6 – 33) (Supplementary Table 1 and Supplementary Fig 1). Raised total cholesterol had the highest code occurrence of unique code sequences (5,408,007) and hypertension had the highest code occurrence including multiple code sequences (29,299,147) (Supplementary Table 2).

### Disease embeddings

We generated two different disease embeddings (see Methods and model pipeline Fig 1 for details of our analysis pipeline). First, we used MCA, a dimensionality reduction technique similar to PCA but specific for binary or categorical data, which extracts components based on occurrence patterns. As shown by the scree plot (Supplementary Fig 2), the first two dimensions explained a large amount of the variance: 58.3% for the first, and 5.5% for the second. As expected, the first dimension largely reflected increasing age and number of conditions (see Supplementary Fig 3). To evaluate our embeddings, three co-authors with clinical expertise developed a set of 253 clinically well-established disease pairs (further details in Methods). Using this set of disease pairs, we retained 30 dimensions from MCA as the number that optimised known disease pairs being assigned in the top ten nearest neighbours to each disease based on the cosine similarity calculated from the MCA embeddings (see Methods and Fig 2).

**Figure 1:**
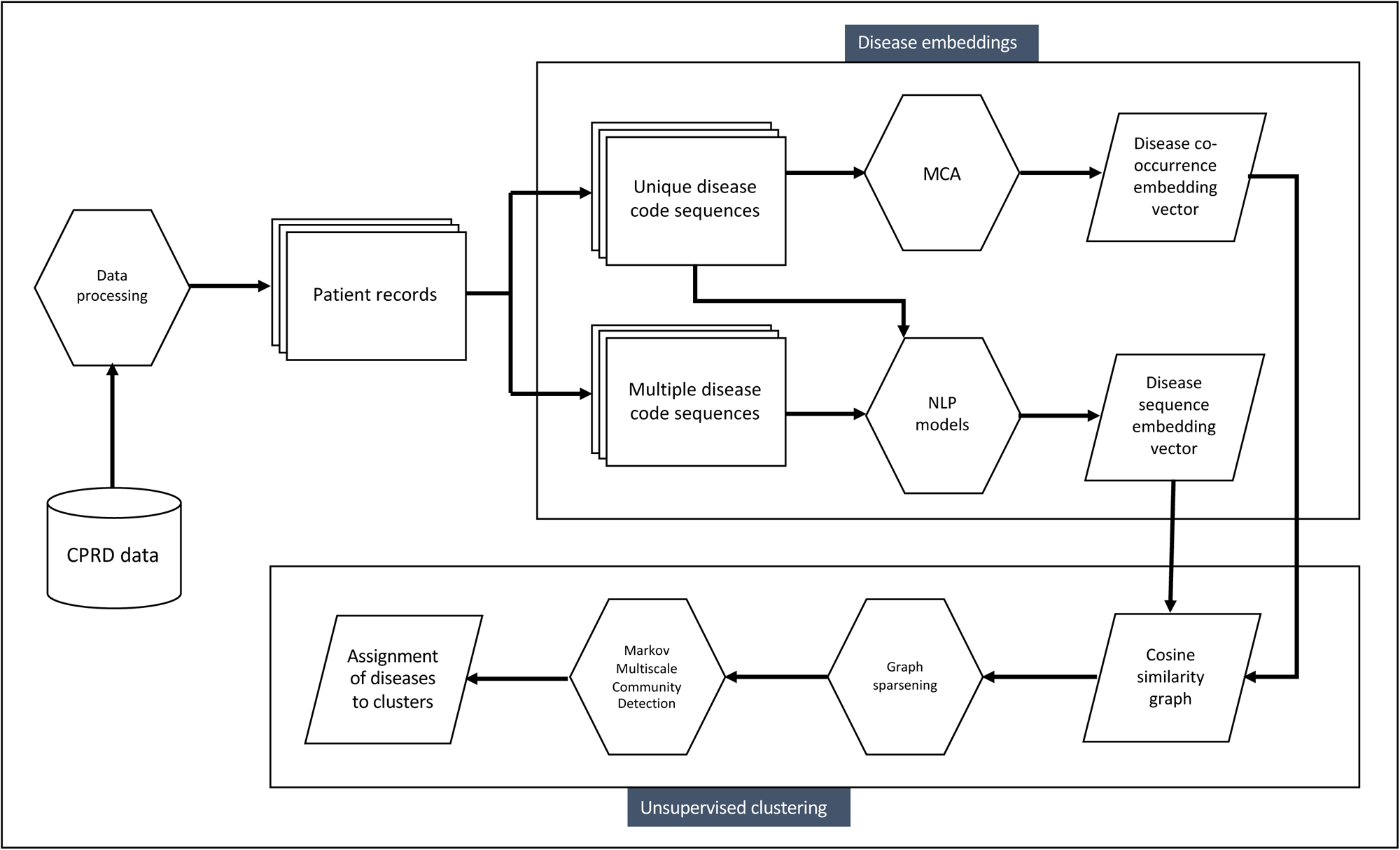
Pipeline for generating disease clusters from Clinical Practice Research Datalink (CPRD) data.

**Figure 2:**
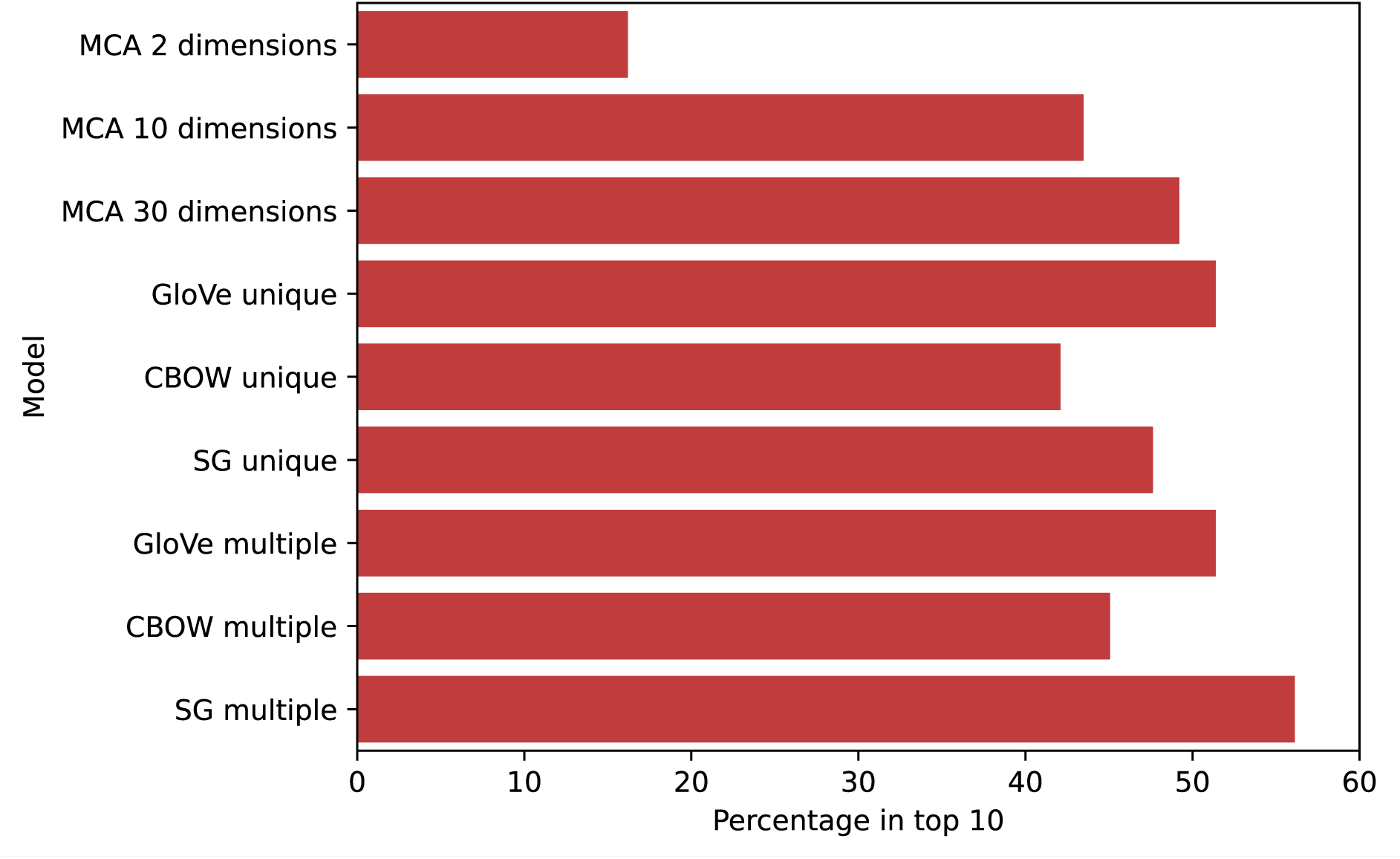
Percentage of disease associations from a curated set of 253 known disease pairs that are assigned to the ten nearest neighbours based on cosine similarity for each disease embedding.

We next generated embeddings using three NLP models: two Word2Vec models using continuous-bag-of-words (CBOW) and skip-gram (SG), and Global Vectors (GloVe). Models were trained on each of the unique and multiple code sequences from all 10.5 million patients. We tested a range of hyperparameter values, and the optimal hyperparameters were chosen for each of the three NLP models using the same evaluation strategy as for MCA (see Supplementary Tables 3-6). When evaluated against the curated set of 253 known disease pairs, GloVe and SG had similar performance to MCA-30 for unique code sequences, with lower performance for CBOW (Fig 2). The NLP models had comparatively better performance when run on multiple code sequences, indicating that additional information is provided by the sequence of reappearing codes. In a sensitivity analysis, model performance was similar when comparing against the nearest two, five or twenty neighbours (Supplementary Fig 4). Overall, SG with multiple codes (SG-M) showed the best performance across all models. We thus selected SG-M with an embedding dimension of 30 as the best-performing NLP embedding and compared it to the best co-occurrence embedding (MCA-30) for clustering.

### Clustering of disease embeddings

We applied a multiscale clustering algorithm (MMCD) to the similarity between both disease embeddings (MCA-30 and SG-M), following the procedure detailed in Methods and Fig 1. Briefly, we first determined the cosine similarity of each disease to each other disease from the disease embeddings. We then used the continuous k-nearest neighbours (CkNN) algorithm to sparsen the similarity matrix. To the resulting similarity graph, we applied the MMCD algorithm to identify a multi-scale sequence of clusterings of increasing coarseness and selected optimal partitions at different levels of resolution. Using the MCA-30 embeddings, MMCD identified optimal clusterings at three resolutions representing 23, nine, and six clusters (Fig 3). Using the SG-M embeddings, optimal clusterings were identified at 25, fifteen, seven, and five clusters and we selected the first three of these for further evaluation (Fig 4). In both cases, Sankey diagrams demonstrated that most conditions in a cluster remained in the same cluster across levels of resolution (Figs 5 and 6). This indicates a quasi-hierarchical pattern of similarity between diseases, with smaller groups of diseases showing greater similarity and, in turn, getting integrated into broader disease groups with a looser observational similarity.

**Figure 3:**
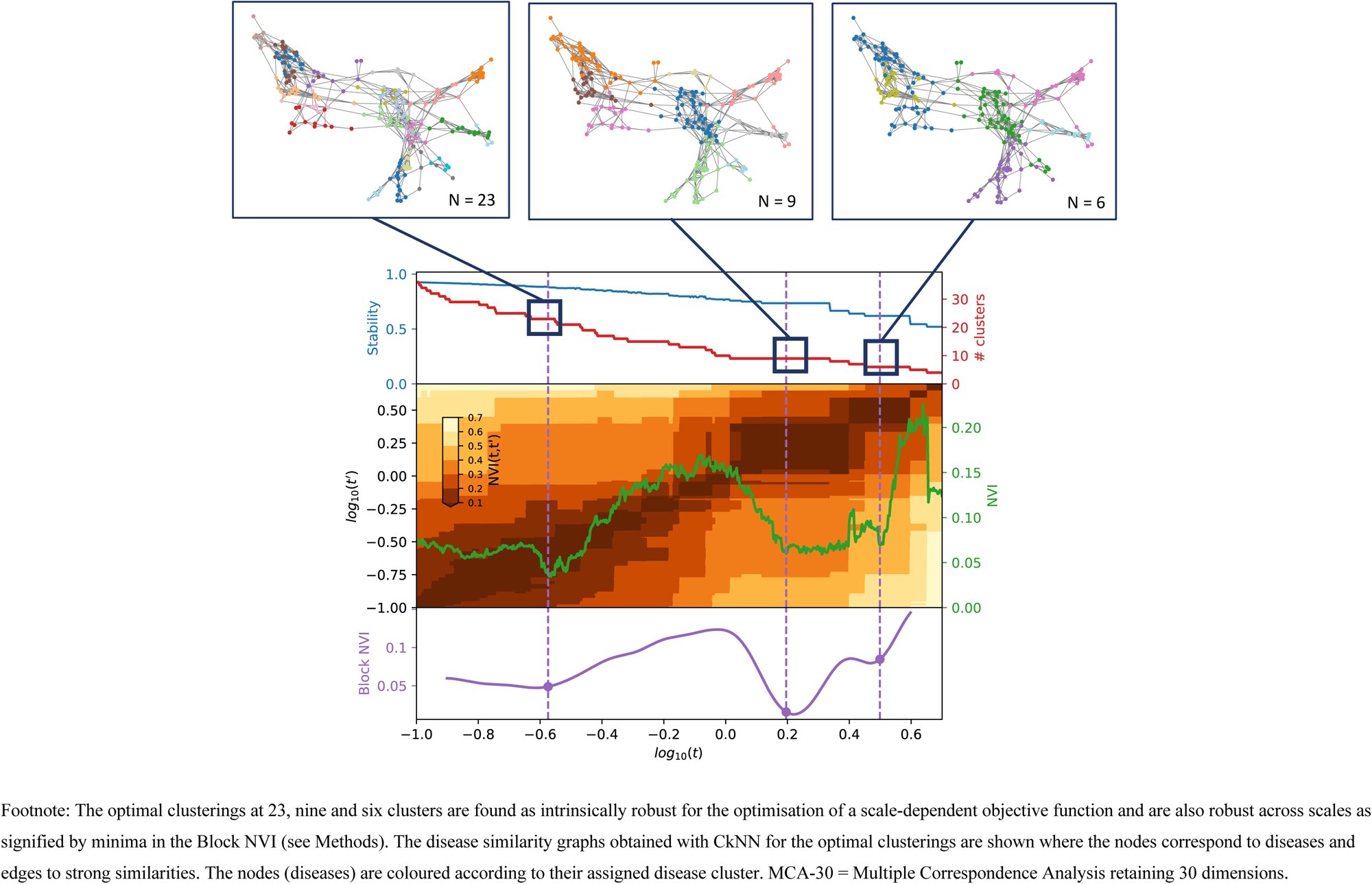
Selection of optimal clusterings from Markov Multiscale Community Detection using MCA-30 embeddings.

**Figure 4:**
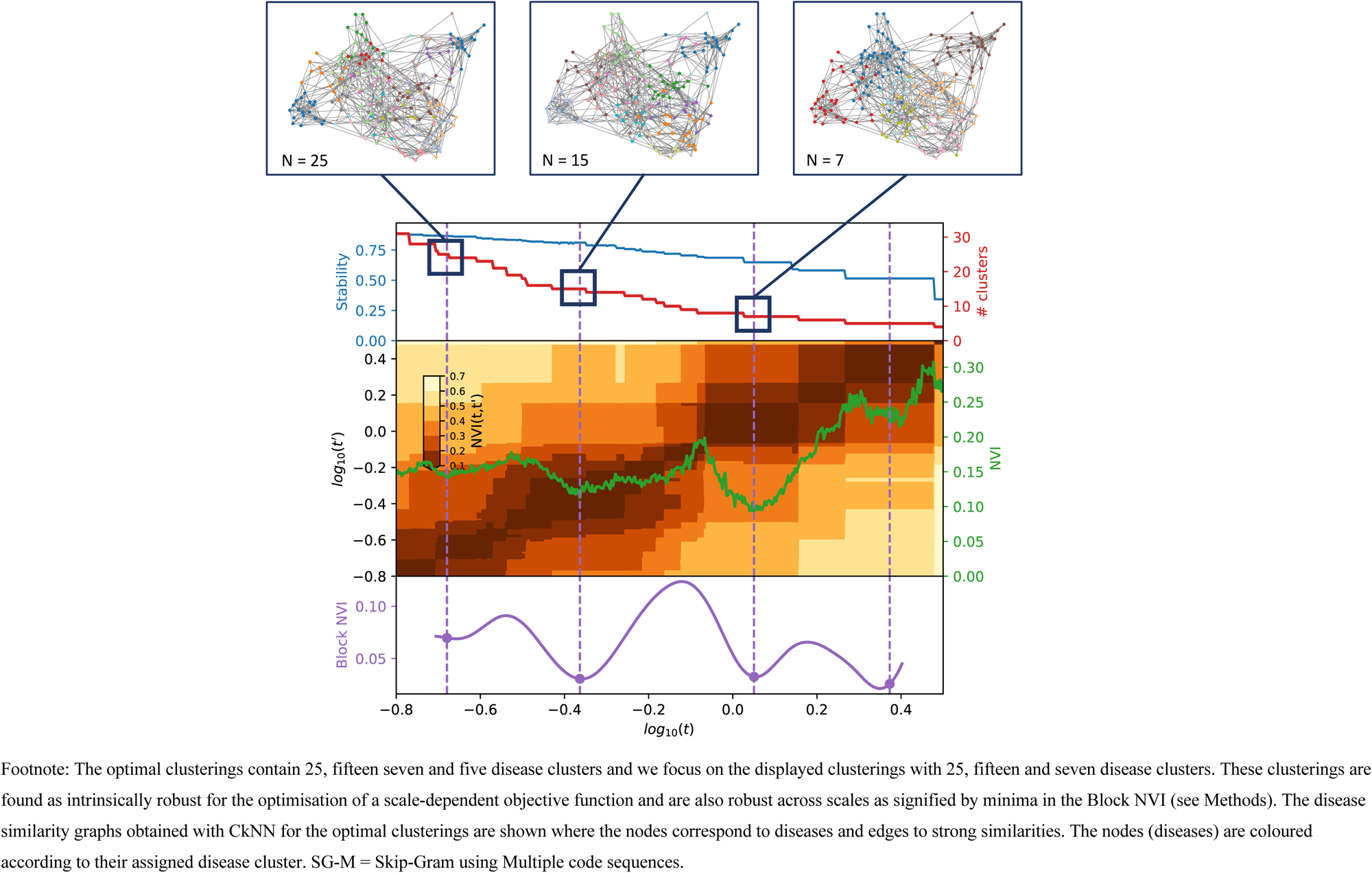
Selection of optimal partitions from Markov Multiscale Community Detection using SG-M embeddings.

**Figure 5:**
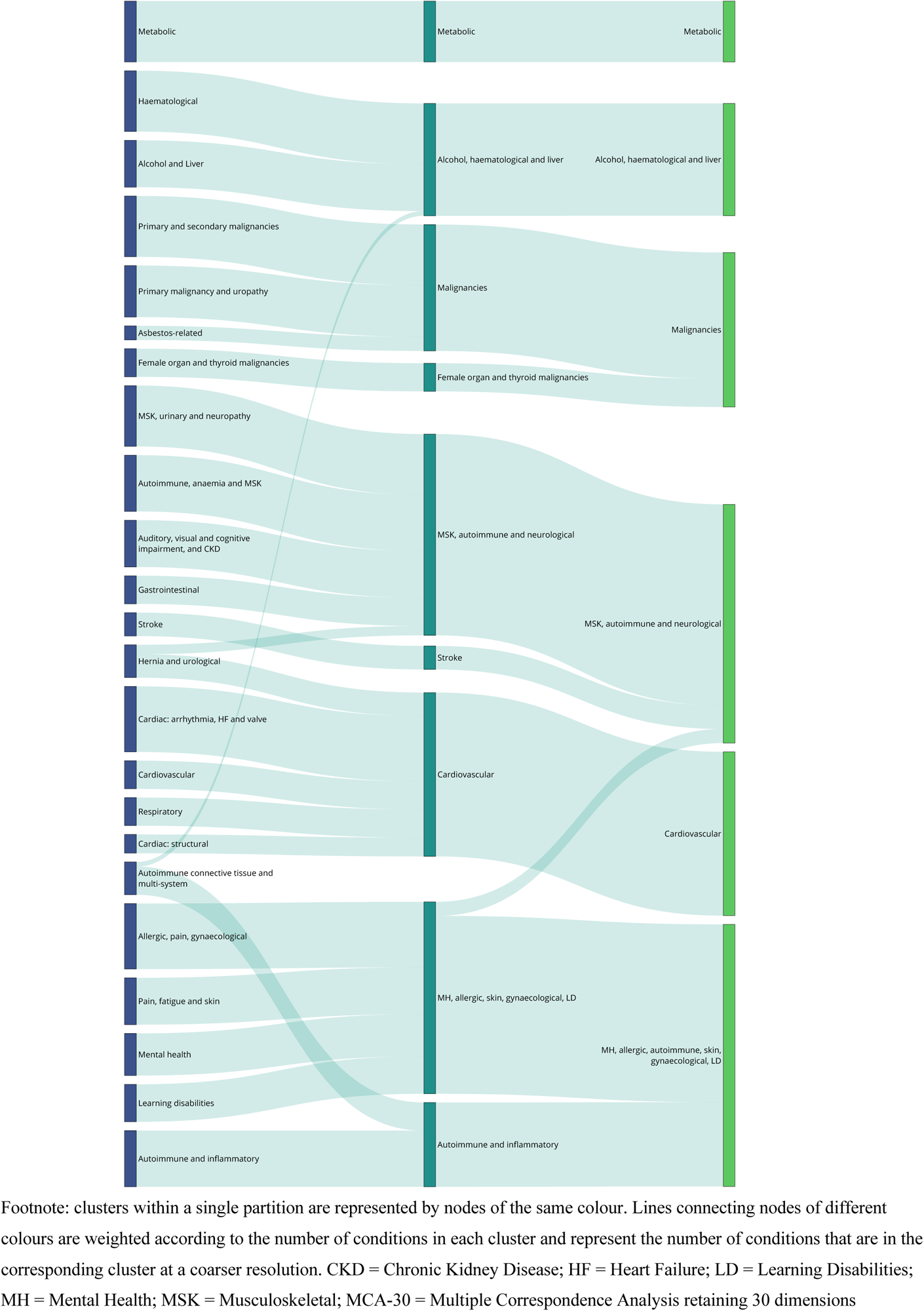
Sankey diagram of clusters at resolutions of 23, 9 and 6 clusters, using MCA-30 embeddings.

**Figure 6:**
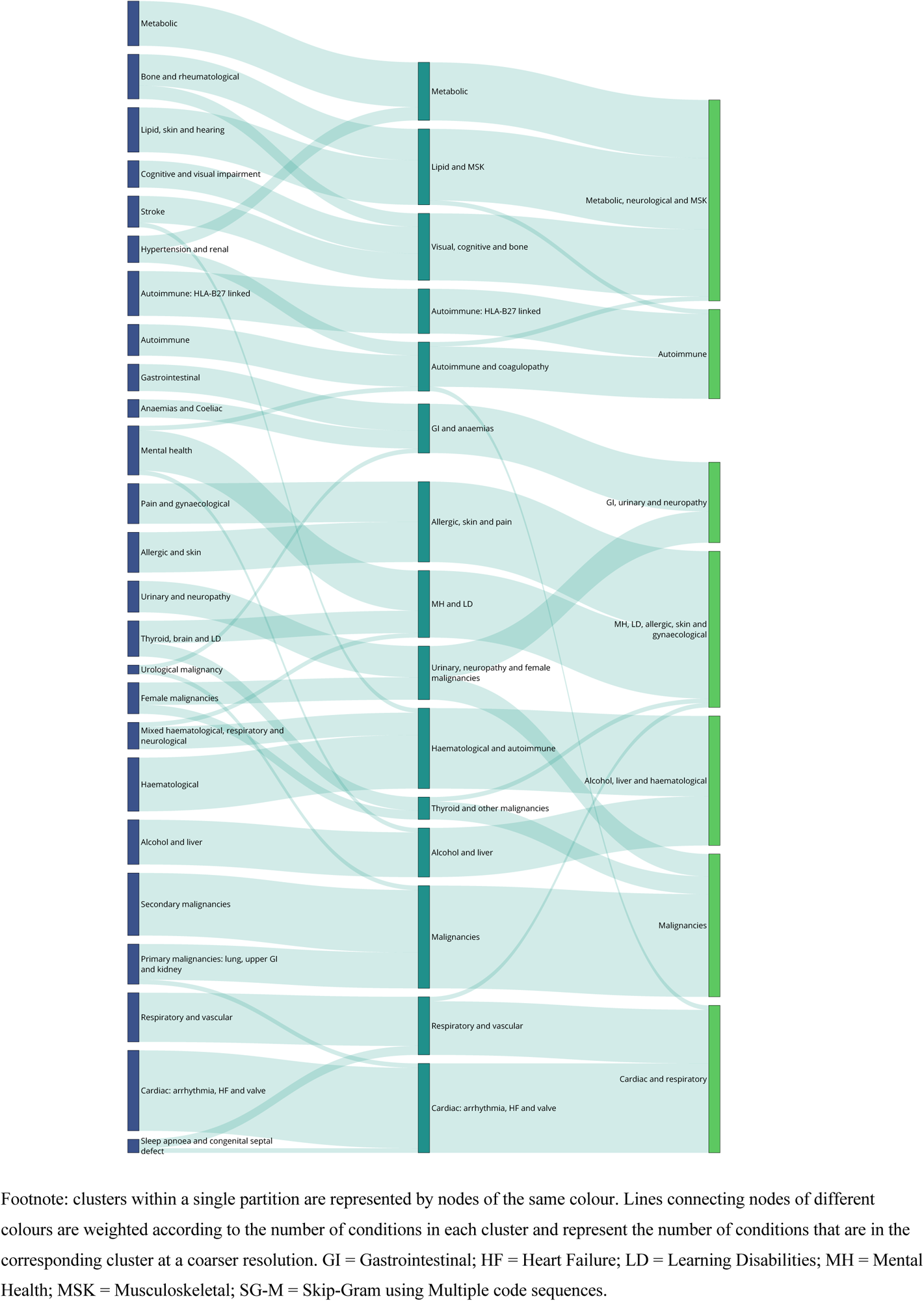
Sankey diagram of clusters at resolutions of 25, 15 and 7 clusters, using SG-M embeddings.

### Comparison to other clustering methods

To evaluate our clustering method, we compared the clusters derived from MMCD to two clustering algorithms widely used in studies of disease clustering: k-means and Ward’s hierarchical clustering.^13^ In each case, we selected the same corresponding number of clusters to those from MMCD. For both embedding methods, k-means and hierarchical clustering produced unbalanced partitions, with a few over-dominant clusters and some additional very small clusters containing few diseases. Using our curated set of 253 clinically established disease associations, we found that known disease pairs were substantially more likely to be assigned to the same cluster using MMCD (Fig 7). Furthermore, although randomly sampling any two diseases from one patient, a patient was more likely to have both conditions assigned to the same disease cluster using hierarchical and k-means clustering, due to the large size of the dominant clusters, they were less likely to share conditions with other people in the same cluster (Supplementary Fig 5) across the range of partitions.

**Figure 7:**
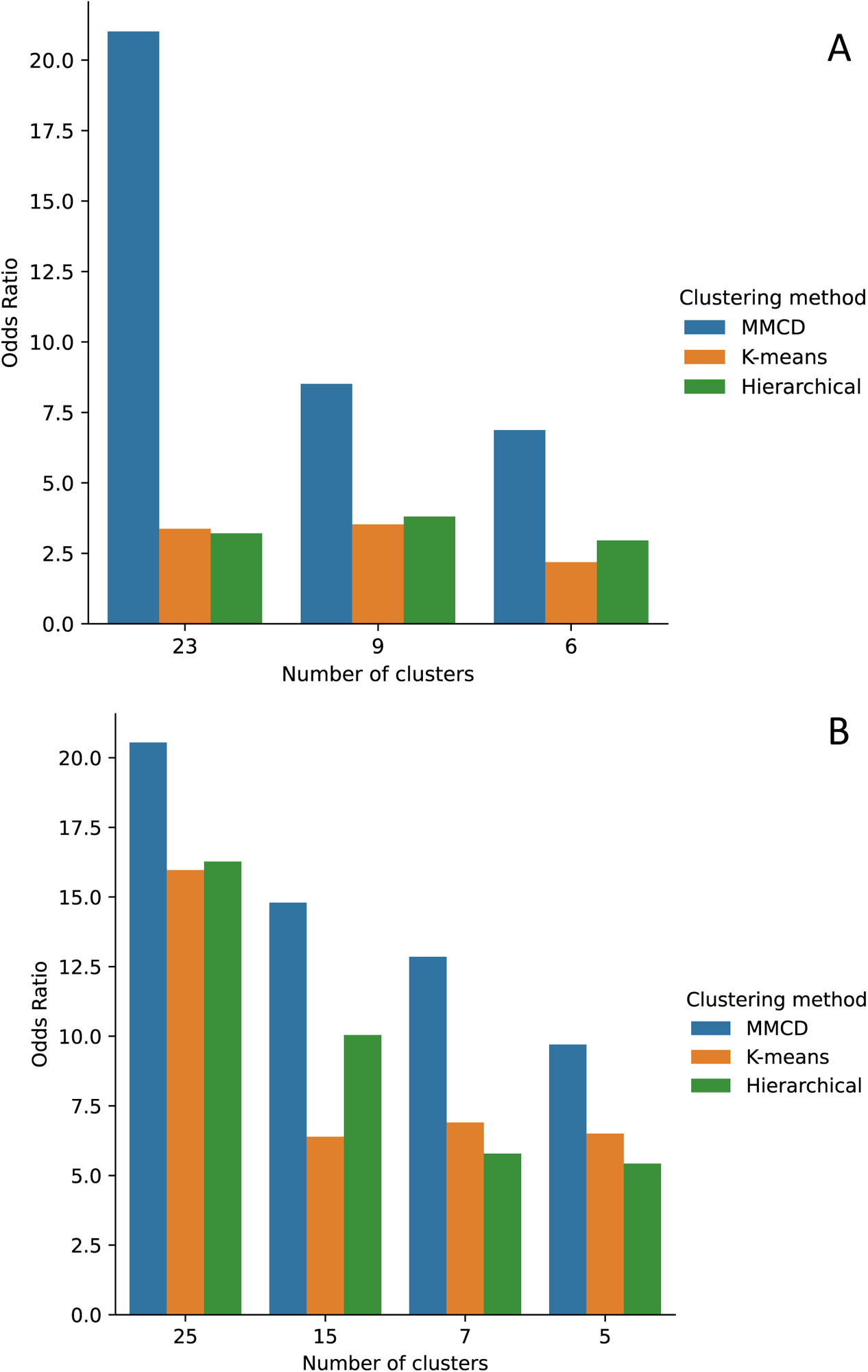
Odds ratios for assigning a known disease pair to the same cluster compared to the expected distribution of 253 known disease pairs, for MCA-30 embeddings (panel A) and SG-M embeddings (panel B)

### Comparison of clusters to ICD-10 chapters

We also compared the MMCD disease clusters to the assignment of the diseases in the corresponding sixteen chapters of the ICD-10 medical taxonomy (see Methods for details) by computing the normalised variation of information (NVI), where NVI=0 indicates perfect agreement and NVI=1 corresponds to maximum disagreement. With the MCA-30 embeddings, the similarity to the ICD-10 chapters ranged from NVI=0.60 for 23 clusters to NVI=0.75 for six clusters (Supplementary Table 7A). With the SG-M embeddings, the NVI was slightly lower than that for MCA-30, ranging from 0.55 for 25 clusters to 0.68 for seven clusters (Supplementary Table 7B). These results indicate a substantial mismatch in the groupings of diseases within the MMCD clusters compared to the ICD-10 chapters, reflecting the difference between data-driven co-occurrence patterns and a clinical taxonomy.

### Descriptive evaluation of clusters

Given its higher performance, we considered only the MMCD clusterings for further descriptive evaluation. To aid visualisation and interpretation, clusters were assigned a descriptive label aiming to represent most of the diseases in the cluster. Figs 5 and 6 show Sankey diagrams capturing the quasi-hierarchical organisation of the MMCD clusters obtained for both MCA-30 and SG-M embeddings, whereas Figs 8 and 9 provide a more detailed visualisation of the contents of the disease clusters.

**Figure 8:**
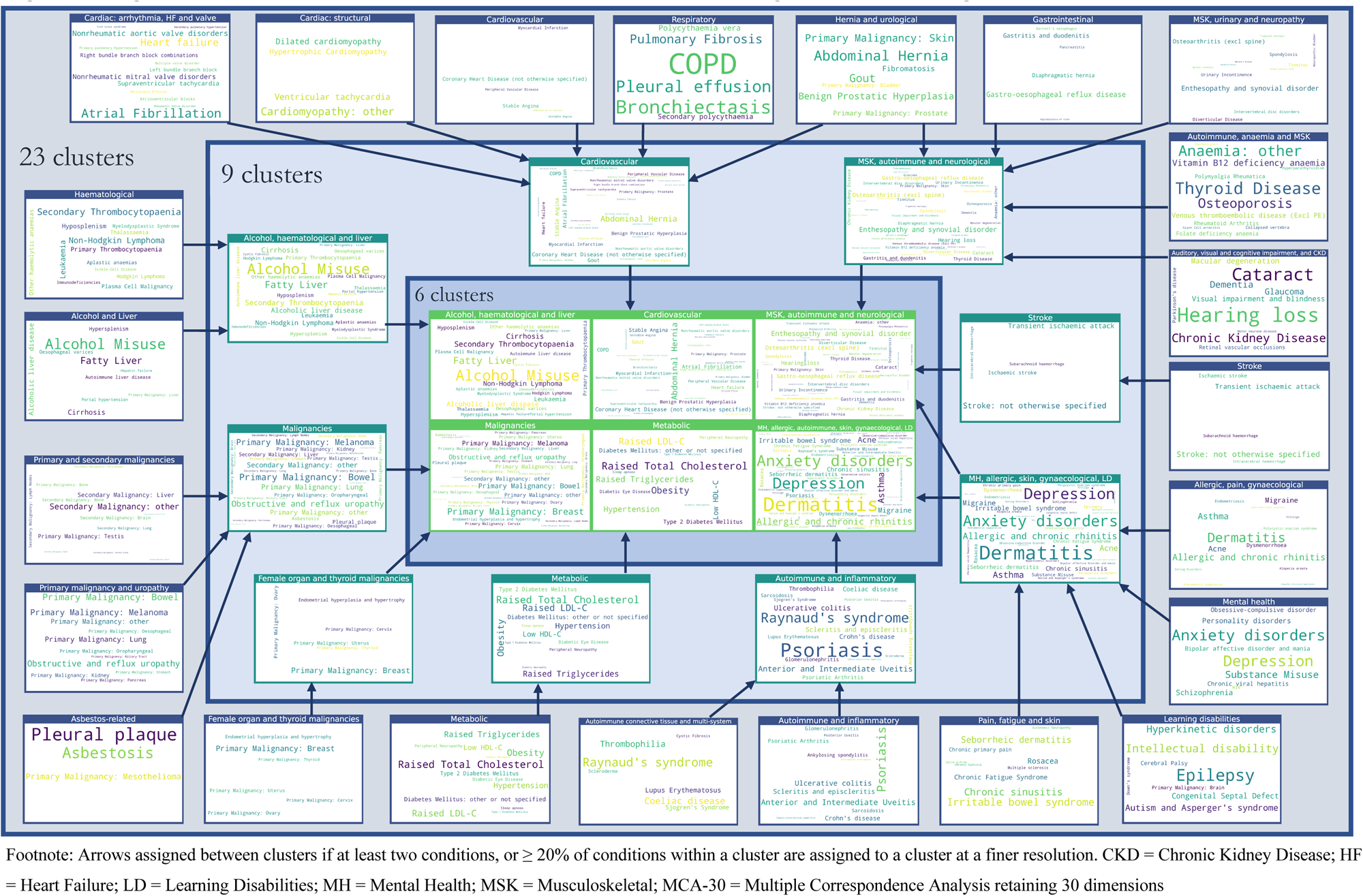
Assignment of diseases to clusters at resolutions of 23, 9 and 6 clusters, using MCA-30 embeddings.

**Figure 9:**
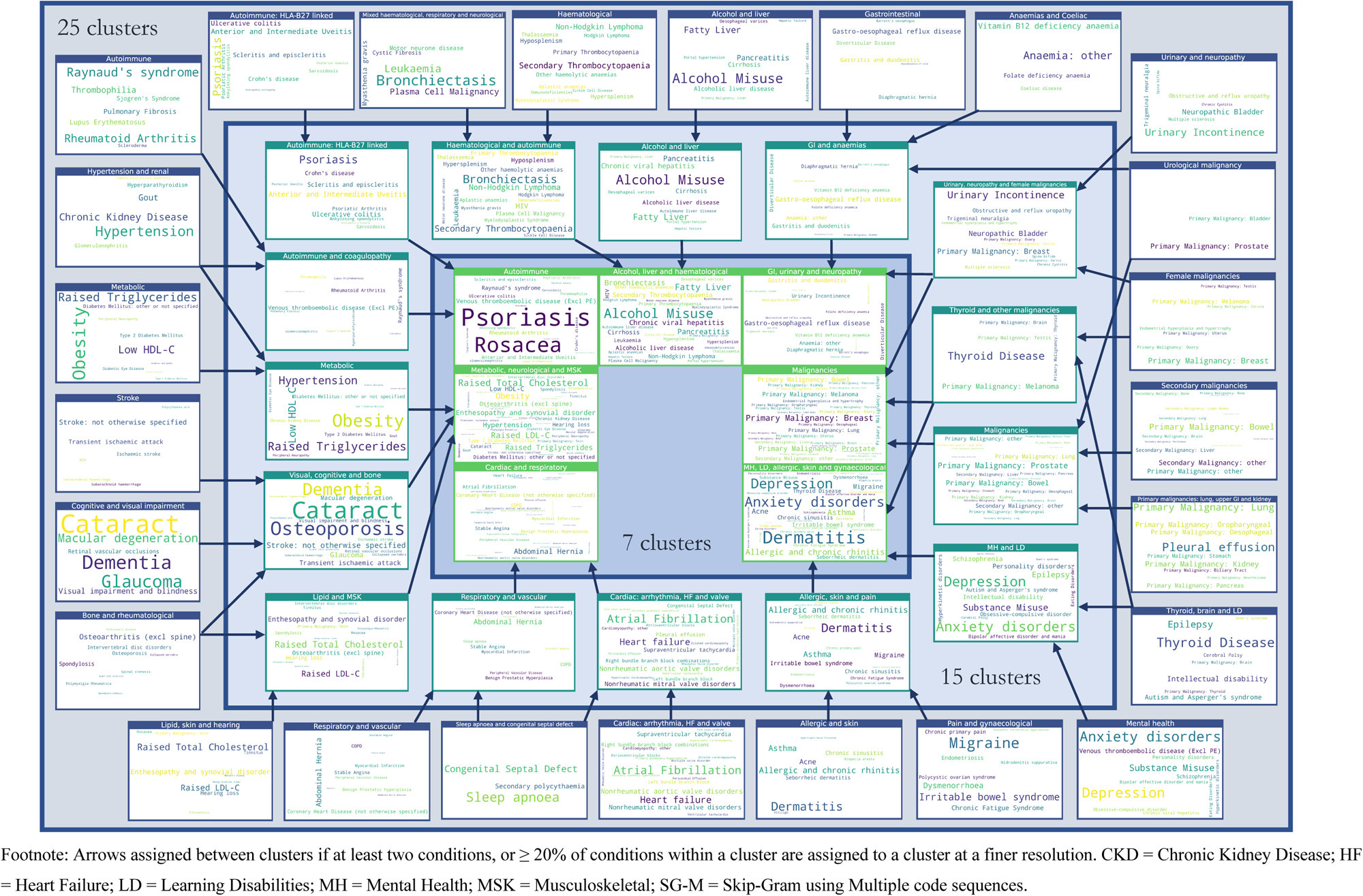
Assignment of diseases to clusters at resolutions of 25, 15 and 7 clusters, using SG-M embeddings.

### 1. Clusters from MCA-30 embeddings

At the 23-cluster resolution, several well-defined clusters were identified, including a cluster representing the established metabolic syndrome^26^ (including obesity, raised cholesterol, hypertension, diabetes and diabetes complications), forms of stroke, autoimmune and inflammatory conditions, and liver conditions (Fig 8). Many malignancies clustered together at this fine resolution, except for breast, gynaecological and thyroid primary malignancies, which clustered separately, and primary malignancy of the skin and prostate, which clustered separately along with urological conditions. As would be expected from similarities drawn from co-occurrences in data, some clusters reflected diseases common in particular age groups, for example a cluster of diseases affecting younger people (including acne, dysmenorrhoea, polycystic ovarian syndrome and allergic and chronic rhinitis), and another cluster with diseases more common in older people (including dementia, hearing loss and visual impairment).

The sequence of clusterings at multiple resolutions revealed that most of the disease clusters at the 23-cluster resolution integrate quasi-hierarchically at coarser resolutions (Fig 5). Notably, the metabolic cluster displayed the strongest stability, with the same conditions clustered together across all resolutions. However, some diseases separated from given clusters across different scales. For example, cystic fibrosis (CF) was present in an ‘Autoimmune and inflammatory’ cluster at a resolution of 23 clusters but joined an ‘Alcohol, haematological and liver’ cluster at coarser resolutions, which may reflect the challenge of assigning a multi-system disease such as CF to a consistent set of clusters.

### 2. Clusters from SG-M embeddings

The clusters derived from the SG-M embeddings, which consider not only co-occurrence patterns but also information contained in the multiple code sequence, were different to those from MCA-30. At the fine 25-cluster resolution, well-defined clusters include, for example, one representing stroke sub-types and another representing heart failure, valvular and arrhythmogenic cardiac conditions (Fig 9). As with MCA-30, a metabolic cluster including diabetes and obesity was observed, but hypertension was clustered instead with renal diseases, and both Raised Total Cholesterol and Raised LDL-C clustered separately with enthesopathy, hearing loss and skin cancer. We found instances of clustering according to underlying causal mechanisms: two separate autoimmune clusters are present in this clustering, one including rheumatoid arthritis and related diseases, and another including spondyloarthropathies and inflammatory bowel disease which are strongly associated with the HLA-B27 gene.^27^

The quasi-hierarchy of the partitions across resolutions in SG-M is less strong than for MCA-30 (Fig 6), reflecting the additional complexities contained in the contextual information of sequences captured by NLP embeddings. For example, thyroid cancer was clustered with thyroid disease and learning disabilities at the 25-cluster resolution, which could be partially attributed to a well-established link between thyroid disease and Down’s syndrome.^28^ At the fifteen-cluster resolution, thyroid disease and thyroid cancer joined a cluster with melanoma, testicular and brain cancer. While people with melanoma may be at higher risk of developing thyroid malignancy^29^, other associations within this cluster were unexpected. To investigate this cluster further, we compared the observed to expected ratio of co-occurrence for each pair of conditions in the cluster (Supplementary Table 8). This demonstrated a stronger than expected ratio of co-occurrence of thyroid disease and thyroid malignancy (9.29), of primary brain cancer with melanoma (2.29), and of thyroid cancer with melanoma (2.35). Testicular cancer had a lower ratio of co-occurrence with thyroid disease and thyroid cancer (0.45 and 0.89, respectively), but higher-than-expected ratio of co-occurrence with brain cancer and melanoma (1.96 and 1.17, respectively), demonstrating that appearance together in a cluster needs to be examined in more detail, as it does not necessarily indicate that each disease is directly associated with every other in the cluster.

## Discussion

Our study presents a novel application of an unsupervised, multiscale graph-based clustering method (MMCD) to vector embeddings of diseases derived from EHR data of 10.5 million patient records. Our analysis produces interpretable clusters of diseases from fine to coarse resolutions, based on the intrinsic patterns of co-occurrence and sequences in people with multimorbidity. We found that MMCD outperformed k-means and hierarchical algorithms in clustering pairs of diseases known to be associated using disease embeddings generated from both co-occurrence-based MCA and sequence-based NLP methods. We also find optimal clusterings over multiple resolutions, highlighting the advantages of considering a range of levels of coarseness. Although a full description of the relationships of all 212 diseases was outside the scope of this study, we demonstrate the power of these methods for classifying multimorbidity clusters at different resolutions, which may help identify more fine-grained relationships in future research. We also provide access to the disease embeddings and cluster assignments, as an open resource for other researchers.

### Clinical implications

Clusters derived from MCA-30 and SG-M embeddings differed, but both pick up meaningful patterns of diseases that are clinically interpretable. In general, clusters from SG-M were less interpretable than those from MCA-30, which is likely to reflect the additional contextual information captured in disease sequences, beyond those captured by co-occurrence alone. It may also reflect differences in coding frequency between diseases, with previous work indicating that some diseases are more likely to have recurrent codes, particularly those with financial incentives attached to their management.[Beaney *et al* (2022), Determinants of disease code frequency in the primary care electronic healthcare record: a retrospective cohort study. Under review at BMJ Open] Although there is no ‘gold-standard’ ground truth for disease clusters to be compared to, conditions that are known to form part of the well-established metabolic syndrome^26^ clustered together across resolutions in MCA-30, while other clusters represented conditions with similar underlying causal mechanisms, for example, those associated with the HLA-B27 gene.^27^

Finer resolutions with more clusters are likely to be the most valuable in identifying novel disease associations and, at these resolutions, we found more unexpected disease patterns that suggest avenues for further investigation, for example, the grouping of thyroid cancer, thyroid disease, melanoma, testicular and brain cancer in SG-M. However, as discussed earlier, it is important to recognise that the cluster assignment does not necessarily indicate that each disease is directly associated with every other disease (Supplementary Table 8). This may be due to a distinguishing feature of sequence-based models compared to co-occurrence-based models, whereby the representation is determined not only by direct co-occurrence but by shared associations of two diseases with other diseases, capturing indirect information which is less obvious.

The clusters demonstrate a remarkably hierarchical structure with MCA-30, and to a lesser extent, with SG-M. It is important to remark that this is an intrinsic feature of the data, rather than MMCD, which does not impose any hierarchical structure on the sequence of clusterings. Our findings suggest that using hierarchical clustering algorithms that enforce a hierarchical structure may mask meaningful variation in the structure of the data over different resolutions. For example, in both MCA-30 and SG-M embeddings, CF appeared with different diseases across resolutions. As a condition linked to both liver disease^30^ and higher prevalence of anxiety and depression,^31^ its separation at different resolutions likely represents the challenge of assigning a multi-system disease to a single branch of hierarchical clusters. Our multi-resolution approach thus provides the advantage of allowing assessment of the stability of disease assignment to clusters across resolutions as a means of drawing further information.

Some studies have evaluated the quality of embeddings by comparing clusters to known hierarchical disease taxonomies, such as ICD-10, which are predominantly based around organ systems rather than aetiology.^19, 20^ We found that our disease clusters were substantially different to the classification of the ICD-10 chapters, highlighting the disparity with systems-based classifications and suggesting that hierarchical taxonomies may not be a suitable method by which to evaluate the quality of disease similarity based on co-occurrence or sequence.

### Implications for embedding and clustering methods

A strength of our work is the direct comparison of co-occurrence to sequence-based embedding methods. With MCA, although practitioners often retain two dimensions to visualise relationships, we demonstrated here that this fails to explain a substantial amount of known disease associations and limits interpretability when using a large set of LTCs. We trialled a range of popular word embedding methods and given the applications of these methods in healthcare data are still relatively new, hypothesised that optimal hyperparameters for text data might not be optimal for disease code sequences, which do not follow the same syntactic relationships. We therefore experimented with a range of hyperparameters and found optimal ranges outside of the default values for standard text applications (see Supplementary Tables 3-6). When using unique code occurrences, we found that both GloVe and SG performed similarly to MCA-30 in identifying known associations, whilst CBOW’s poorer performance was in line with previous reports in both text and healthcare data.^32, 33^ That NLP models and MCA produce similar results when using unique code sequences is unsurprising given the common basis in using disease-disease co-occurrence. Where NLP models showed substantial improvement (see SG-M in Fig 2) was when using longer sequences that included recurrent codes in the record, thus utilising additional information beyond direct co-occurrence. Previous studies have shown that sequence-based models have superior predictive performance for a range of outcomes,^18, 34^ but we additionally found that the generated embeddings also better reflect clinically known disease associations.

To help alleviate the lack of a gold-standard set of disease clusters, we created a list of established disease pairs and used these to compare across methods, finding MMCD to perform substantially better than both k-means and hierarchical clustering, particularly at finer resolutions. K-means and hierarchical clustering both produced unbalanced clusters with a large, dominant cluster and other smaller clusters of rarely occurring diseases. The effect was more marked for MCA-30, suggesting a smoothing effect of the SG-M-generated embeddings when compared to MCA. However, in both cases, MMCD produced more balanced clusters, likely due to both the sparsification of the network using the MST-CkNN algorithm and the clustering cost function (Markov Stability), which enables MMCD to overcome such problems when using highly dimensional data.

### Future work

In future, we plan to extend our methods to cluster patients directly, using approaches analogous to topic modelling and document embeddings in NLP,^35, 36^ specifically using large language models, such as BERT, which may provide additional insights into the similarity of disease sequences across people.^20, 34^ Although previous studies have evaluated the association of disease clusters with patient outcomes^9, 37^ we believe that evaluating outcomes should be reserved for clusters of patients, rather than clusters of diseases. Indeed, our preliminary assessment showed that disease clusters are not directly representative of patients, as relatively few patients were allocated to a single disease cluster, even when only two of their diseases were randomly sampled. Various approaches have been used to assign patients to disease clusters based, e.g., on a patient having one or more,^38^ two or more,^39, 40^ or three or more^41^ diseases in a cluster. However, these methods can assign patients to multiple clusters, an issue that will escalate with a larger number of clusters, and which can bias assessment of outcomes and complicate clinical use.

With MCA, age was a strong contributor to the first dimension which explained the largest proportion of variance. Furthermore, some of our clusters reflected sex differences, such as the clustering of gynaecological and breast malignancies at finer resolutions. Future work could consider stratification of clustering by age and sex, which may increase the ability to detect associations between less common diseases. Although our study already used a larger number of diseases (212) than previous studies of multimorbidity disease clustering, further increasing the number of conditions or using individual diagnostic codes (rather than categorising into diseases) may also increase the ability to detect novel associations at finer resolutions.

### Strengths and limitations

A strength of our study is the use of a large and representative sample of patients registered to general practices in England which enhances the generalisability of our results.^42^ We used a larger set of LTCs than used previously in multimorbidity research to aid replicability, which we have made available to other researchers. We experimented with a range of hyperparameters for our NLP algorithms and selected the best performing models. Nevertheless, it is possible that better-performing models exist outside the range of hyperparameters tested.

To compare embedding models, we developed a list of clinically-established disease associations. However, this list is not exhaustive, and may be biased towards inclusion of more common conditions which have a stronger evidence base. Furthermore, the combination of conditions included in the CALIBER study may also lead to bias in the embeddings and the clustering. For example, several unique conditions describe forms of liver disease and its sequelae (alcoholic liver disease, hepatic failure, cirrhosis, portal hypertension, oesophageal varices), which may all represent the same pathophysiological process, and so may inflate similarity metrics between these conditions. This may explain the prominent separation of liver diseases on the second dimension in MCA (Supplementary Fig 3). However, other authors using different data sources and definitions have found similarly strong clustering of liver-related conditions.^24^ Similarly, the stability of the metabolic cluster across resolutions in MCA-30 may in part stem from the inclusion of more diseases of this type in the code-lists (five diseases representing diabetes and its complications, and four representing cholesterol and triglycerides).

There are examples where a disease may be classified as both a specific and non-specific version of the same disease, both of which may appear in a patient’s record. For example, codes for ‘Diabetes: other or not specified’ may be found in a patient’s record in addition to those for ‘Type 2 diabetes’ or ‘Type 1 diabetes’, and similarly ‘Stroke: not otherwise specified’ in addition to ‘Ischaemic stroke’ or ‘Intracerebral haemorrhage’. In these cases, the non-specific disease may represent use of generic codes used across each disease subtype, rather than that the disease itself is ‘other’ or unspecified’ and are likely to be explained by clinician coding practices and the specificity of the available codes. As a result, the embeddings and clusters generated from routinely collected EHR data as used here reflect not only disease co-occurrence, but factors related to patients, clinicians, and healthcare organisations.[Beaney *et al* (2022), Determinants of disease code frequency in the primary care electronic healthcare record: a retrospective cohort study. Under review at BMJ Open]

## Conclusion

In conclusion, using a representative cohort of over ten million people registered to general practices in England, we found clusters of diseases corresponding to both established and novel patterns. Clusters derived from co-occurrence-based embedding methods tended to be more straightforward to interpret than those from sequence-based NLP embedding methods, likely reflecting the additional relationships captured in disease sequences. Our multi-resolution approach highlights the nearly hierarchical structure of disease clusters but with notable exceptions that indicate the complexity of categorising certain diseases into a single set of inclusive clusters. Our study demonstrates the promise of these methods for identifying patterns of disease clusters within highly dimensional healthcare data, which could be used to facilitate discovery of associations between diseases in the future and help in optimising the management of people with multimorbidity, which is a priority for health systems globally.

## Methods

### Data sources and data cleaning

We used the CPRD Aurum dataset, a nationally representative source of general practice data in England.^42^ We included all patients aged 18 years or over, registered to a GP practice in CPRD Aurum between 1^st^ January 2015 and 1^st^ January 2020. Patients were censored at the earliest of date of deregistration, date of death, date of last collected data extraction from the practice, or the 1^st^ January 2020. Any codes that were recorded or observed after the censoring date were excluded (see Supplementary Methods for details). Patients with two or more of the diseases defined below were included. Data cleaning rules for variables, including socio-demographics, are explained in detail in the Supplementary Methods.

### Disease definitions

Diagnostic codes are recorded in CPRD as Medcodes. These are entered by clinicians during clinical consultations and converted into a numeric code, for example, the term ‘Allergic asthma’ as Medcode ID 1483199016. We translated codes to a corresponding set of 212 LTCs based on code-lists developed for the CALIBER study, from which 211 conditions were selected relevant to LTCs by Head *et al* (2021). For example, the diagnostic codes representing ‘Allergic asthma’ and ‘Exercise induced asthma’ are grouped under the disease category of ‘asthma’. We reviewed these codes and supplemented them with an additional disease of chronic primary pain as a prevalent condition often included in multimorbidity studies (see Supplementary Methods).^45, 46^ Diseases were ordered in sequences from earliest to latest according to timestamp of the observation, for example, a patient record might read sequentially as: “asthma, asthma, type 2 diabetes, hypertension, asthma, hypertension”. We constructed two sequences for comparison: the first (“multiple”) used all codes, and the second (“unique”) included a disease only at its first occurrence (i.e., date of diagnosis); in the example above, this sequence is simplified as: “asthma, type 2 diabetes, hypertension”. Where two codes had the same timestamp, we randomly ordered the corresponding codes.

## Statistical methods

### Generating disease embeddings

Fig 1 summarises the steps of our pipeline from data processing to clustering. We compared four different methods to create disease embeddings. As a baseline approach used previously in multimorbidity research, we used MCA.^24, 25^ Correspondence analysis (CA) is a class of methods which aim to reduce the dimensionality of binary or categorical data, analogous to Principal Component Analysis for continuous data, by minimising the chi-squared distance between observed and expected values based on the global co-occurrence matrix, or *Burt* matrix.^47–49^ MCA is an extension of CA to two or more variables and has an advantage of allowing supplementary variables to be added which do not contribute to the calculation of the variance.^48^ We applied MCA to the disease co-occurrence matrix, using the MCA algorithm implemented in Stata version 17.0 (StataCorp).^50^

We compared MCA to three popular NLP word embedding models: the Word2vec models using CBOW and SG,^32^ and GloVe.^51^ CBOW and SG are related methods which use neural network architectures: in the case of CBOW, the model predicts a target word given a surrounding context window, whereas in SG, the model predicts the context given a target word.^32^ In contrast, GloVe incorporates matrix factorisation of global co-occurrence statistics, combined with a local window.^51^ In each case, we compared the default hyperparameter values of the models to values we hypothesised might better represent the smaller vocabulary and relatively short sequences (in comparison to the documents for which the methods were originally developed). We then selected the best performing model according to our evaluation metrics below.

For CBOW and SG, we used the Word2vec model implemented in the *gensim* package for training on the sequences of all 10.5 million patients.^52^ We compared vector sizes of 10 and 30, window sizes of 2 and 5, negative sampling of 2 and 5 and down-sampling of frequent diseases comparing the default of 0.001 to no down-sampling. For GloVe, we used the *glove-python* implementation and compared a default window size of 5 to values of 2, and learning rate of 0.05 to values of 0.01 and 0.1.^53^ We also tested models over a range of epochs as detailed in the Supplementary Methods.

### Evaluation of embeddings

We evaluated our embedding methods using a curated set of 253 known disease association pairs. These were created by three co-authors with a clinical background, TB, JC and DS, based on the 212 available diseases, using the British Medical Journal Best Practice guidelines and clinical judgement as detailed in the Supplementary Methods.^54^ For each embedding model we proceeded as follows: for each disease *d_1_* in the set of known disease association pairs we calculated the percentage of known associated diseases (*d_2_*_…_ *d_N_*) that were in the set of ten most similar diseases of *d_1_* in terms of cosine similarity computed from the embedding. Similar approaches have been used by other authors, with Solares *et al* (2021) using a range of neighbourhood sizes from three to 20.^20^ Beam *et al* (2019) used bootstrap sampling of the similarity distribution for each condition, and assigned conditions if in the top 5% of the distribution of most similar conditions, which is roughly equivalent to use of the top ten conditions in our case (given 212 conditions).^22^ We checked the robustness of our evaluation by comparing different thresholds of neighbourhood sizes, and found they produced similar optimal models (Supplementary Fig 4).

### Markov Multiscale Community Detection

To cluster the selected disease embeddings, we used MMCD. The first step is to construct a similarity graph of diseases, a sparsified weighted graph where the diseases are the nodes of the graph and the weights represent the similarity between the embedding vectors. To construct the graph from the data, we calculated the pairwise cosine similarity matrix *S* for all diseases and followed the normalisation approach of Altuncu *et al* (2019), by calculating the distance matrix *D* = 1 − *S*, applying max normalisation to give 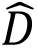, and then calculating the normalised cosine similarity as 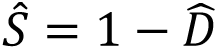.^55^ This produces a dense similarity matrix, which is then sparsified to transform it into a *similarity graph*. This sparsification is a key pre-processing step in MMCD as it removes edges with weaker associations. Although simple thresholding based on weights was originally applied in this step, it is not robust to noise and does not capture well the inhomogeneities in the similarities in the data. Hence, several methods have been proposed for this step using global constructions that involve the minimum spanning tree (MST), which contains the collection of edges with minimum weight sum that fully connect all nodes on the graph, thus ensuring global connectivity. To this sparse network, edges representing local connectivity are often added, such as the k-nearest neighbours (kNN) for each node. Recently, Liu and Barahona (2020) demonstrated improvement on the kNN graph by using continuous kNN (CkNN).^17^ In CkNN, for distance *d_i,j_* connecting nodes *i* and *j*, and where *d^k^(i)* and *d^k^(j)* are the distances to the *k*-th nearest neighbour of *i* and *j*, respectively, then the edge is retained if:

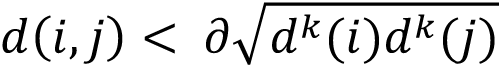

where ∂ is a parameter that can be varied to alter the sparsity of the network. In our case, we hold ∂ constant at a value of 1, but vary the value of *k*; as shown by Liu and Barahona (2020), the MMCD algorithm is relatively robust to different parameterisations.^17^ We selected a CkNN value of ten, but comparison of CkNN values of five, fifteen and twenty resulted in similar partitions.

To this sparsened undirected network, we then applied MMCD, using the *pygenstability* module in Python^16, 56, 57^ which models a random-walk across the network, and evaluates subgraphs of the original graph over which the Markov dynamics is contained over a time (*t)* that acts as a scale. The natural scanning over scales performed by the diffusion on the graph reveals larger communities (i.e., coarser clusterings) as the scale *t* is increased.^16^ For details of the method and its applications see Delvenne *et al* (2010) and Arnaudon *et al* (2021).^16, 56^ At each time step, we optimise the cost function 2,000 times using the Leiden algorithm,^58^ and calculate the normalised variation of information (NVI), an information theoretic measure for comparing cluster partitions, where 0 indicates identical partitions and 1 indicates dissimilar partitions. The algorithm then selects partitions that have low values of the NVI across scales and also with respect to the Leiden optimisation, using the automated scale selection algorithm developed by Schindler *et al* (2023) which smooths the NVI to identify persistence across scales.^60^ Models were run over a Markov scale aiming for between 4 and 30 clusters, using 500 scale steps, 2000 optimisation evaluations and select 400 optimisations to compute the NVI at each scale.^57^

### Benchmarking and evaluation of clusters

As a benchmark, we compared the cluster partitions derived from MMCD to k-means and hierarchical clustering using Ward’s method, as baselines widely used in multimorbidity research.^13^ We compared to the same number of clusters as identified in MMCD. In contrast to MMCD, which used the cosine similarity matrix, these methods were applied directly to the disease embeddings as input features. We implemented k-means with the Lloyd algorithm, iterating 1,000 times with different random centroid seeds. We used *scikit-learn* package for both k-means and Ward’s clustering algorithms.^61^

To enable comparison across methods, we calculated metrics related to the interpretability of clusters. As an intrinsic measure of the relevance of disease clusters to patterns of diseases in patients, we first randomly sampled 100,000 patients with replacement. For each patient, we then randomly sampled two different diseases from their set of all diseases, once per patient. We assigned patients to a disease cluster if *both* diseases were contained within the same cluster. Of these patients {*P_1_,… P_N_*} assigned to a disease cluster, we calculated a metric of the pairwise Jaccard similarity between the set of two diseases {*d_1_, d_2_*} for each patient in the same cluster, and report the arithmetic mean of all possible pairs.

To compare between the three clustering algorithms for partitions with the same number of clusters, we used information from the 253 known disease pairs. We expect that in a more interpretable clustering solution, known disease pairs are more likely to be assigned to the same cluster. To correct for the bias that favours unbalanced and uninformative clustering solutions with all diseases assigned to a single cluster, we considered the observed assignment of known disease pair edges within clusters to that expected, assuming the contingency table in Table 1.

**Table 1:** contingency table of assignment of known disease pairs to clusters.

|  | Intra-cluster | Inter-cluster |
| --- | --- | --- |
| Known disease pair edges | $DP_{intra}$ | $DP_{inter}$ |
| Other disease pair edges | $E_{intra}$ | $E_{inter}$ |

Following from Table 1, we calculated the odds ratio for a known disease pair edge being intra-cluster compared to inter-cluster:

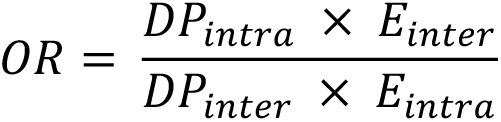

A higher OR here can be interpreted as higher odds that a known disease pair edge is found in the same cluster given the cluster distribution for a partition with the given number of clusters, representative of more balanced and informative clusters.

### Comparison to ICD-10 classification

We compared the clusters to the ‘system’ a condition is assigned to in the CALIBER code-lists, which is corresponds closely to the classification of chapters in ICD-10,^43^ using the NVI. Each disease is assigned to one of sixteen ‘systems’, for example, ‘asthma’ is assigned under ‘Diseases of the Respiratory System’, similar to the chapters used in ICD-10.

We used Python version 3.8.10 and Pandas version 1.3.5 for data manipulation and management.^62, 63^ Sankey diagrams were created in Plotly.^64^

## Data Availability

This study uses patient level which is not publicly available but can be requested for users meeting certain requirements: https://cprd.com/research-applications. The code lists and embeddings generated from this work are available to download from: https://tbeaney.github.io/MMclustering/.

## Acknowledgements

This research is funded through a clinical PhD fellowship awarded to TB from the Wellcome Trust 4i programme at Imperial College London. Data management was provided by the Big Data and Analytical Unit (BDAU) at the Institute of Global Health Innovation (IGHI), Imperial College London. We are grateful for the support of the NIHR Imperial Biomedical Research Centre. JC acknowledges support from the Wellcome Trust (215938/Z/19/Z). DS is supported by an Imperial College and National Institute of Health Research (NIHR) Post-Doctoral, Post-CCT research fellowship. TW, AM and PA acknowledge support from the National Institute for Health and Care Research (NIHR) Applied Research Collaboration Northwest London. MB acknowledges support from EPSRC grant EP/N014529/1 supporting the EPSRC Centre for Mathematics of Precision Healthcare. We thank Dominik Schindler for helpful discussions and help with the use of the PyGenStability package. The views expressed in this publication are those of the authors and not necessarily those of the NHS, the NIHR, the Wellcome Trust or the Department of Health and Social Care.

## Ethical approval

Data access to the Clinical Practice Research Datalink (CPRD) and ethical approval was granted by CPRD’s Research Data Governance Process on 28^th^ April 2022 (Protocol reference: 22_001818).

## References

1. The Academy of Medical Sciences. Multimorbidity: a priority for global health research. Academy of Medical Sciences (2018).

2. Pearson-Stuttard, J., Ezzati, M. & Gregg, E. W. Multimorbidity—a defining challenge for health systems. The Lancet Public Health 4, e599–e600 (2019).

3. Makovski, T. T., Schmitz, S., Zeegers, M. P., Stranges, S. & van den Akker, M. Multimorbidity and quality of life: Systematic literature review and meta-analysis. Ageing Research Reviews 53, 100903 (2019).

4. Nunes, B. P., Flores, T. R., Mielke, G. I., Thumé, E. & Facchini, L. A. Multimorbidity and mortality in older adults: A systematic review and meta-analysis. Arch Gerontol Geriatr 67, 130–138 (2016).

5. Soley-Bori, M. et al. Impact of multimorbidity on healthcare costs and utilisation: a systematic review of the UK literature. British Journal of General Practice 71, e39--e46 (2021).

6. Zulman, D. M. et al. Multimorbidity and healthcare utilisation among high-cost patients in the US Veterans Affairs Health Care System. BMJ Open 5, (2015).

7. Jani, B. D. et al. Relationship between multimorbidity, demographic factors and mortality: findings from the UK Biobank cohort. BMC Medicine 17, 74 (2019).

8. Vetrano, D. L. et al. Trajectories of functional decline in older adults with neuropsychiatric and cardiovascular multimorbidity: A Swedish cohort study. PLOS Medicine 15, e1002503 (2018).

9. Jackson, C. A. et al. Multimorbidity patterns are differentially associated with functional ability and decline in a longitudinal cohort of older women. Age Ageing 44, 810–816 (2015).

10. BMJ Best Practice. Metabolic syndrome - Symptoms, diagnosis and treatment. https://bestpractice.bmj.com/topics/en-gb/212.

11. Kassi, E., Pervanidou, P., Kaltsas, G. & Chrousos, G. Metabolic syndrome: definitions and controversies. BMC Medicine 9, 48 (2011).

12. Whitty, C. J. M. & Watt, F. M. Map clusters of diseases to tackle multimorbidity. Nature 579, 494–496 (2020).

13. Busija, L., Lim, K., Szoeke, C., Sanders, K. M. & McCabe, M. P. Do replicable profiles of multimorbidity exist? Systematic review and synthesis. European Journal of Epidemiology 34, 1025–1053 (2019).

14. Prados-Torres, A., Calderón-Larrañaga, A., Hancco-Saavedra, J., Poblador-Plou, B. & van den Akker, M. Multimorbidity patterns: a systematic review. Journal of Clinical Epidemiology 67, 254–266 (2014).

15. Schaub, M. T., Lehmann, J., Yaliraki, S. N. & Barahona, M. Structure of complex networks: Quantifying edge-to-edge relations by failure-induced flow redistribution. Network Science 2, 1–24 (2014).

16. Delvenne, J.-C., Yaliraki, S. N. & Barahona, M. Stability of graph communities across time scales. Proceedings of the National Academy of Sciences of the United States of America 107, 12755–60 (2010).

17. Liu, Z. & Barahona, M. Graph-based data clustering via multiscale community detection. Appl Netw Sci 5, 1–20 (2020).

18. Rasmy, L., Xiang, Y., Xie, Z., Tao, C. & Zhi, D. Med-BERT: pretrained contextualized embeddings on large-scale structured electronic health records for disease prediction. *npj Digit*. Med. 4, 1–13 (2021).

19. Choi, E. et al. Multi-layer representation learning for medical concepts. Proceedings of the ACM SIGKDD International Conference on Knowledge Discovery and Data Mining 13-17-Augu, 1495–1504 (2016).

20. Solares, J. R. A. et al. Transfer Learning in Electronic Health Records through Clinical Concept Embedding. 1–14 (2021).

21. Choi, E., Schuetz, A., Stewart, W. F. & Sun, J. Medical Concept Representation Learning from Electronic Health Records and its Application on Heart Failure Prediction. Preprint at http://arxiv.org/abs/1602.03686 (2017).

22. Beam, A. L. et al. Clinical Concept Embeddings Learned from Massive Sources of Multimodal Medical Data. Preprint at http://arxiv.org/abs/1804.01486 (2019).

23. Sourial, N. et al. Correspondence analysis is a useful tool to uncover the relationships among categorical variables. Journal of clinical epidemiology 63, 638–646 (2010).

24. Bisquera, A. et al. Identifying longitudinal clusters of multimorbidity in an urban setting: A population-based cross-sectional study. The Lancet Regional Health - Europe 3, 100047 (2021).

25. Guisado-Clavero, M. et al. Multimorbidity patterns in the elderly: a prospective cohort study with cluster analysis. BMC Geriatrics 18, 16 (2018).

26. Cornier, M.-A. et al. The Metabolic Syndrome. Endocr Rev 29, 777–822 (2008).

27. Parameswaran, P. & Lucke, M. HLA B27 Syndromes. in StatPearls (StatPearls Publishing, 2023).

28. Karlsson, B., Gustafsson, J., Hedov, G., Ivarsson, S.-A. & Annerén, G. Thyroid dysfunction in Down’s syndrome: relation to age and thyroid autoimmunity. Archives of Disease in Childhood 79, 242–245 (1998).

29. Zerfaoui, M. et al. New Insights into the Link between Melanoma and Thyroid Cancer: Role of Nucleocytoplasmic Trafficking. Cells 10, 367 (2021).

30. Kobelska-Dubiel, N., Klincewicz, B. & Cichy, W. Liver disease in cystic fibrosis. Przegl^a̜^ d Gastroenterologiczny 9, 136 (2014).

31. Guta, M. T. et al. Global Burden of Anxiety and Depression among Cystic Fibrosis Patient: Systematic Review and Meta-Analysis. Int J Chronic Dis 2021, 6708865 (2021).

32. Mikolov, T., Chen, K., Corrado, G. & Dean, J. Efficient estimation of word representations in vector space. 1st International Conference on Learning Representations, ICLR 2013 - Workshop Track Proceedings 1–12 (2013).

33. Dynomant, E. et al. Word Embedding for the French Natural Language in Health Care: Comparative Study. JMIR Medical Informatics 7, e12310 (2019).

34. Li, Y. et al. BEHRT: Transformer for Electronic Health Records. Sci Rep 10, 7155 (2020).

35. Altuncu, M. T., Yaliraki, S. N. & Barahona, M. Graph-Based Topic Extraction from Vector Embeddings of Text Documents: Application to a Corpus of News Articles. Studies in Computational Intelligence 944, 154–166 (2021).

36. Lannou, E. L., et al. Clustering of patient comorbidities within electronic medical records enables high-precision COVID-19 mortality prediction. medRxiv 2021.03.29.21254579 (2021).

37. Vetrano, D. L. et al. Twelve-year clinical trajectories of multimorbidity in a population of older adults. Nature Communications 11, 3223 (2020).

38. Foguet-Boreu, Q. et al. Multimorbidity patterns in elderly primary health care patients in a South Mediterranean European region: A cluster analysis. PLoS ONE 10, 1–14 (2015).

39. Prados-Torres, A. et al. Multimorbidity Patterns in Primary Care: Interactions among Chronic Diseases Using Factor Analysis. PLOS ONE 7, e32190 (2012).

40. Kirchberger, I. et al. Patterns of Multimorbidity in the Aged Population. Results from the KORA-Age Study. PLOS ONE 7, e30556 (2012).

41. Schäfer, I. et al. Multimorbidity Patterns in the Elderly: A New Approach of Disease Clustering Identifies Complex Interrelations between Chronic Conditions. PLOS ONE 5, e15941 (2010).

42. Wolf, A. et al. Data resource profile: Clinical Practice Research Datalink (CPRD) Aurum. International Journal of Epidemiology 48, 1740–1740g (2019).

43. Kuan, V. et al. A chronological map of 308 physical and mental health conditions from 4 million individuals in the English National Health Service. The Lancet Digital Health 1, e63–e77 (2019).

44. Head, A. et al. Inequalities in incident and prevalent multimorbidity in England, 2004&#x2013;19: a population-based, descriptive study. The Lancet Healthy Longevity 2, e489–e497 (2021).

45. Bisquera, A. et al. Inequalities in developing multimorbidity over time: A population-based cohort study from an urban, multi-ethnic borough in the United Kingdom. Lancet Reg Health Eur 12, 100247 (2021).

46. Ashworth, M. et al. Journey to multimorbidity: longitudinal analysis exploring cardiovascular risk factors and sociodemographic determinants in an urban setting. BMJ Open 9, (2019).

47. Hair, J. F., Black, W. C., Babin, B. J. & Anderson, R. E. Multivariate Data Analysis. (Pearson Education Limited, 2013).

48. Michael Greenacre. Correspondence Analysis in Practice. (Chapman & Hall/CRC, 2007).

49. Greenacre, M. TYING UP THE LOOSE ENDS IN SIMPLE, MULTIPLE AND JOINT CORRESPONDENCE ANALYSIS.

50. StataCorp. Stata 17 Base Reference Manual. (Stata Press, 2021).

51. Pennington, J., Richard, S. & Manning, C. D. GloVe: Global Vectors for Word Representation. EMNLP (2014).

52. Řehůřek, R. & Sojka, P. Software Framework for Topic Modelling with Large Corpora. in Proceedings of the LREC 2010 Workshop on New Challenges for NLP Frameworks 45–50 (ELRA, 2010).

53. Kula, M. glove-python. https://github.com/maciejkula/glove-python (2023).

54. British Medical Journal Best Practice. https://bestpractice.bmj.com.

55. Altuncu, M. T., Mayer, E., Yaliraki, S. N. & Barahona, M. From free text to clusters of content in health records: an unsupervised graph partitioning approach. Applied Network Science 4, 2 (2019).

56. Arnaudon, A. et al. PyGenStability: Multiscale community detection with generalized Markov Stability. Preprint at http://arxiv.org/abs/2303.05385 (2023).

57. Barahona Research - Applied Math - Imperial. PyGenStability. https://github.com/barahona-research-group/PyGenStability (2023).

58. Traag, V. A., Waltman, L. & van Eck, N. J. From Louvain to Leiden: guaranteeing well-connected communities. Sci Rep 9, 5233 (2019).

59. Vinh, N. X., Epps, J. & Bailey, J. Information Theoretic Measures for Clusterings Comparison: Variants, Properties, Normalization and Correction for Chance.

60. Schindler, D. J., Clarke, J. & Barahona, M. Multiscale mobility patterns and the restriction of human movement. Preprint at http://arxiv.org/abs/2201.06323 (2023).

61. 2.3. Clustering — scikit-learn 0.24.1 documentation. https://scikit-learn.org/stable/modules/clustering.html#k-means.

62. The Python Language Reference. Python documentation https://docs.python.org/3/reference/index.html.

63. McKinney, W. Data Structures for Statistical Computing in Python. in 56–61 (2010). doi:10.25080/Majora-92bf1922-00a.

64. Plotly Technologies Inc. Collaborative data science. https://plot.ly (2015).

