## Supplementary Appendix for "Identifying multi-resolution clusters of diseases in ten million patients with multimorbidity in primary care in England"

### Long-term conditions (LTCs)

Diagnostic codes were mapped to diseases using code lists developed for the CALIBER study, which have been adapted for use in multimorbidity and CPRD Aurum.[43,44] We reviewed the codes in these lists, and made amendments to the code lists for diabetes, to remove Type 1 and Type 2 codes from the other/unspecified code list. We added chronic primary pain to the set of included conditions and created a new code list. Previous studies of multimorbidity in primary care settings have found a high prevalence and burden of chronic pain.[45,46]

### Curating list of known disease pairs

We created a list of 253 known disease association pairs. These were put together by three co-authors with clinical experience, TB, JC and DS. Each of the 212 LTCs were considered for inclusion, and known associations were created using the British Medical Journal Best Practice guidelines as a source, combined with clinical interpretation. An initial set of associations was developed by TB and each code pair was reviewed and discussed with JC and DS and refined further. We aimed to generate a list of well-established disease associations, and excluded those with a low evidence-based, or those which were non-specific (for example, depression is often listed as a complication of LTCs).

### Table 1: Socio-demographic characteristics of the study population

| Patient characteristic | Total | Percent |
|---|---|---|
| **Age (years)** | | |
| Mean (SD) | 52.2 (20.0) | |
| Median (IQR) | 52 (36 – 68) | |
| 18-29 | 1722749 | 16.3% |
| 30-39 | 1492148 | 14.1% |
| 40-49 | 1754523 | 16.6% |
| 50-59 | 1700192 | 16.1% |
| 60-69 | 1504689 | 14.2% |
| 70-79 | 1265502 | 12.0% |
| 80+ | 1139429 | 10.8% |
| **Gender** | | |
| Female | 5645522 | 53.4% |
| Indeterminate | 263 | <0.1% |
| Male | 4933447 | 46.6% |
| **Ethnicity** | | |
| White | 7727978 | 73.0% |
| South Asian | 684792 | 6.5% |
| Black | 397900 | 3.8% |
| Other | 166501 | 1.6% |
| Mixed | 132468 | 1.3% |
| Missing | 1469593 | 13.9% |
| **IMD* decile** | | |
| 1 (most deprived) | 1107779 | 10.5% |
| 2 | 1042599 | 9.9% |
| 3 | 1080575 | 10.2% |
| 4 | 1089697 | 10.3% |
| 5 | 1007896 | 9.5% |
| 6 | 1067110 | 10.1% |
| 7 | 1090890 | 10.3% |
| 8 | 1069964 | 10.1% |
| 9 | 1053205 | 10.0% |
| 10 (least deprived) | 958965 | 9.1% |
| Missing | 10552 | 0.1% |
| **Number of disease codes per patient** | | |
| Unique codes | | |
| Mean (SD) | 6.51 (4.5) | |
| Median (IQR) | 5 (3 – 9) | |
| Multiple codes | | |
| Mean (SD) | 29.1 (49.4) | |
| Median (IQR) | 13 (6 – 33) | |
| **Total** | **10579232** | |

* IMD = Index of Multiple Deprivation

**Figure 1: Boxplots of distribution of codes per patient for unique versus multiple code sequences**

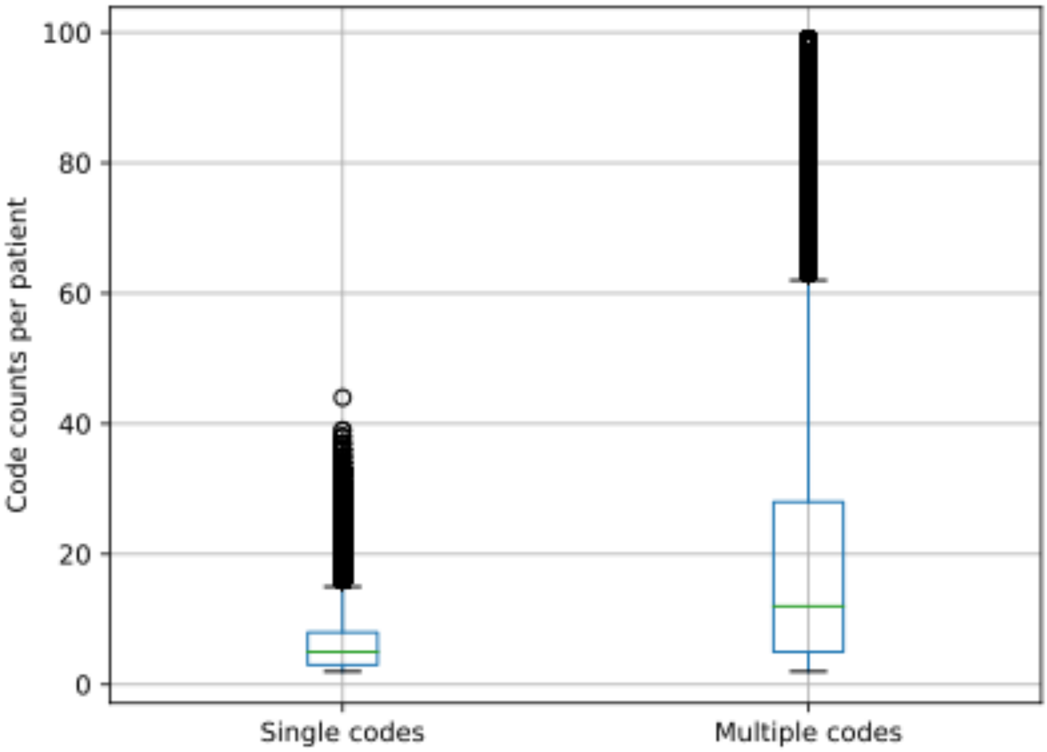


Note: patients with code sequences >100 excluded from figure to aid comparison

**Table 2: Code counts for each disease, when using unique or multiple code sequences**

| Disease | Count including multiple code sequences | Count including unique code sequences |
|---|---|---|
| Hypertension | 29299147 | 2566996 |
| Obesity | 26738855 | 3889266 |
| Raised Total Cholesterol | 21254448 | 5408007 |
| Diabetes Mellitus: other or not specified | 17930916 | 1088793 |
| Depression | 14764407 | 2578030 |
| Type 2 Diabetes Mellitus | 12905910 | 949518 |
| Asthma | 12455656 | 2040019 |
| Raised LDL-C | 11582571 | 4040151 |
| Anxiety disorders | 11044361 | 2588744 |
| Raised Triglycerides | 10372773 | 2931317 |
| Dermatitis | 7717177 | 3220014 |
| Low HDL-C | 7269770 | 2196930 |
| Osteoarthritis (excl spine) | 6149283 | 1594700 |
| Enthesopathy and synovial disorder | 5720143 | 2676007 |
| Thyroid Disease | 5321262 | 751946 |
| COPD | 4581144 | 396214 |
| Allergic and chronic rhinitis | 4492882 | 2128435 |
| Coronary Heart Disease (not otherwise specified) | 4172594 | 522956 |
| Acne | 3546267 | 1482915 |
| Atrial Fibrillation | 3428365 | 477351 |
| Vitamin B12 deficiency anaemia | 2841474 | 360322 |
| Migraine | 2836992 | 1124186 |
| Gastro-oesophageal reflux disease | 2659603 | 1186284 |
| Substance Misuse | 2460092 | 314043 |
| Chronic Kidney Disease | 2380427 | 703739 |
| Epilepsy | 2274387 | 232209 |
| Hearing loss | 2225559 | 1230130 |
| Abdominal Hernia | 2181087 | 960660 |
| Alcohol Misuse | 2162303 | 905096 |
| Psoriasis | 2079122 | 527783 |
| Stable Angina | 2048244 | 418691 |
| Irritable bowel syndrome | 1987218 | 891587 |
| Cataract | 1828115 | 819481 |
| Gout | 1819622 | 446715 |
| Dementia | 1731948 | 308173 |

| | | |
|---|---|---|
| Rheumatoid Arthritis | 1720518 | 119212 |
| Osteoporosis | 1534641 | 422911 |
| Chronic sinusitis | 1391537 | 774486 |
| Heart failure | 1355406 | 262517 |
| Diabetic Eye Disease | 1352166 | 412640 |
| Diverticular Disease | 1330943 | 546711 |
| Gastritis and duodenitis | 1284759 | 755216 |
| Spondylosis | 1283953 | 567301 |
| Urinary Incontinence | 1257820 | 569417 |
| Anaemia: other | 1257406 | 582950 |
| Type 1 Diabetes Mellitus | 1256770 | 79626 |
| Stroke: not otherwise specified | 1206360 | 284057 |
| Diaphragmatic hernia | 1196712 | 510601 |
| Myocardial Infarction | 1192253 | 274607 |
| Schizophrenia | 1148376 | 121606 |
| Glaucoma | 1139439 | 237090 |
| Benign Prostatic Hyperplasia | 1133005 | 317557 |
| Primary Malignancy: Breast | 1067346 | 196856 |
| Chronic Fatigue Syndrome | 1027070 | 244837 |
| Polymyalgia Rheumatica | 982877 | 112479 |
| Seborrheic dermatitis | 978036 | 593671 |
| Intervertebral disc disorders | 977271 | 418824 |
| Primary Malignancy: Skin | 972381 | 381845 |
| Primary Malignancy: Prostate | 953167 | 124710 |
| Rosacea | 842120 | 353502 |
| Tinnitus | 773209 | 476516 |
| Peripheral Vascular Disease | 742257 | 156508 |
| Transient ischaemic attack | 731052 | 200117 |
| Intellectual disability | 685046 | 93963 |
| Dysmenorrhoea | 683534 | 425299 |
| Bipolar affective disorder and mania | 666270 | 66497 |
| Peripheral Neuropathy | 647232 | 305229 |
| Ulcerative colitis | 639792 | 83516 |
| Chronic primary pain | 634565 | 125906 |
| Venous thromboembolic disease (Excl PE) | 590138 | 208586 |
| Crohn's disease | 577766 | 56842 |
| Bronchiectasis | 470411 | 78282 |
| Parkinson's disease | 457306 | 51106 |
| Multiple sclerosis | 435261 | 36334 |
| Personality disorders | 427160 | 111798 |
| Primary Malignancy: Bowel | 413092 | 88252 |
| Macular degeneration | 392186 | 173572 |

| | | |
|---|---|---|
| Neuropathic Bladder | 381732 | 179425 |
| Endometriosis | 369765 | 149537 |
| Fibromatosis | 333800 | 149817 |
| Nonrheumatic aortic valve disorders | 310466 | 123227 |
| Psoriatic Arthritis | 307737 | 39529 |
| Coeliac disease | 306232 | 56348 |
| Sleep apnoea | 303599 | 159192 |
| Obsessive-compulsive disorder | 295214 | 73994 |
| Spinal stenosis | 283930 | 96146 |
| Visual impairment and blindness | 276175 | 165513 |
| Raynaud's syndrome | 268424 | 160595 |
| Ischaemic stroke | 262845 | 73605 |
| Barrett's oesophagus | 261318 | 75657 |
| Anterior and Intermediate Uveitis | 252470 | 108705 |
| Hyperkinetic disorders | 239081 | 67307 |
| Supraventricular tachycardia | 220132 | 71141 |
| Pancreatitis | 220127 | 78478 |
| Primary Malignancy: Melanoma | 217889 | 73642 |
| Trigeminal neuralgia | 215428 | 68213 |
| Autism and Asperger's syndrome | 211600 | 66932 |
| Polycystic ovarian syndrome | 202658 | 102158 |
| Primary Malignancy: Lung | 198855 | 46747 |
| Nonrheumatic mitral valve disorders | 193623 | 101391 |
| Fatty Liver | 188023 | 129813 |
| Eating Disorders | 185196 | 62384 |
| Non-Hodgkin Lymphoma | 182496 | 37713 |
| Chronic viral hepatitis | 180366 | 68204 |
| Lupus Erythematosus | 174633 | 23093 |
| Folate deficiency anaemia | 172387 | 122701 |
| Unstable Angina | 171174 | 81083 |
| Cirrhosis | 168159 | 44633 |
| Hidradenitis suppurativa | 165282 | 58930 |
| Primary Malignancy: Bladder | 161912 | 39012 |
| Retinal vascular occlusions | 158206 | 65224 |
| Meniere's Disease | 147148 | 42593 |
| Ankylosing spondylitis | 146578 | 24988 |
| Leukaemia | 145118 | 27463 |
| Giant Cell arteritis | 144602 | 22595 |
| Secondary Malignancy: other | 136672 | 44026 |
| Primary Malignancy: other | 134340 | 39221 |
| Glomerulonephritis | 128601 | 31738 |
| Diabetic Neuropathy | 125467 | 49135 |

| | | |
|---|---|---|
| Alcoholic liver disease | 124724 | 39213 |
| Obstructive and reflux uropathy | 123707 | 73849 |
| Sarcoidosis | 122670 | 29707 |
| Alopecia areata | 120010 | 83387 |
| Cardiomyopathy: other | 115723 | 26421 |
| Scleritis and episcleritis | 103197 | 65745 |
| Collapsed vertebra | 102459 | 48319 |
| Secondary Thrombocytopaenia | 100387 | 50671 |
| Pleural effusion | 100035 | 60843 |
| Benign essential tremor | 99167 | 46059 |
| Plasma Cell Malignancy | 97358 | 12716 |
| Hyperparathyroidism | 96993 | 37320 |
| HIV | 95411 | 28909 |
| Pulmonary Fibrosis | 94002 | 24260 |
| Vitiligo | 91178 | 58191 |
| Congenital Septal Defect | 88317 | 45437 |
| Thrombophilia | 80592 | 47077 |
| Primary Malignancy: Oropharyngeal | 79258 | 14537 |
| Subarachnoid haemorrhage | 77044 | 28692 |
| Cerebral Palsy | 76283 | 17950 |
| Intracerebral haemorrhage | 73679 | 23640 |
| Spondylolisthesis | 72284 | 38292 |
| Primary Malignancy: Ovary | 71977 | 18471 |
| Primary Malignancy: Oesophageal | 71675 | 12922 |
| Autoimmune liver disease | 67481 | 11337 |
| Abdominal Aortic Aneurysm | 61002 | 27922 |
| Sjogren's Syndrome | 59763 | 16045 |
| Primary Malignancy: Brain | 59613 | 11501 |
| Primary Malignancy: Kidney | 58376 | 16751 |
| Primary Malignancy: Uterus | 56829 | 18566 |
| Right bundle branch block combinations | 56673 | 45121 |
| Autonomic Neuropathy | 55869 | 21629 |
| Left bundle branch block | 54000 | 35188 |
| Hyposplenism | 52991 | 18258 |
| Primary Malignancy: Thyroid | 50026 | 9331 |
| Myasthenia gravis | 48537 | 5926 |
| Primary Thrombocytopaenia | 48052 | 17594 |
| Chronic Cystitis | 44829 | 15198 |
| Cystic Fibrosis | 44438 | 3721 |
| Dilated cardiomyopathy | 43434 | 11035 |
| Hodgkin Lymphoma | 42511 | 10450 |
| Polycythaemia vera | 41335 | 12183 |

| | | |
|---|---|---|
| Primary Malignancy: Testis | 40686 | 12782 |
| Primary Malignancy: Cervix | 40077 | 13536 |
| Primary Malignancy: Pancreas | 39440 | 8951 |
| Myelodysplastic Syndrome | 37362 | 8456 |
| Spina bifida | 37095 | 15910 |
| Other haemolytic anaemias | 36353 | 14471 |
| Scleroderma | 35985 | 5910 |
| Sickle Cell Disease | 34844 | 4637 |
| Idiopathic Intracranial Hypertension | 34066 | 8281 |
| Hypertrophic Cardiomyopathy | 33974 | 8236 |
| Down's syndrome | 33448 | 7578 |
| Oesophageal varices | 33382 | 12986 |
| Pleural plaque | 32066 | 18482 |
| Secondary Malignancy: Liver | 31564 | 15696 |
| Atrioventricular blocks | 30841 | 22967 |
| Primary Malignancy: Stomach | 29912 | 6400 |
| Asbestosis | 29396 | 13183 |
| Ventricular tachycardia | 28726 | 12501 |
| Motor neurone disease | 28165 | 4528 |
| Rheumatic Valve Disorder | 28126 | 9069 |
| Secondary polycythaemia | 24811 | 11254 |
| Primary pulmonary hypertension | 23353 | 11248 |
| Hypersplenism | 22884 | 15329 |
| Aplastic anaemias | 22203 | 9996 |
| Endometrial hyperplasia and hypertrophy | 20449 | 16030 |
| Thalassaemia | 20343 | 12424 |
| Posterior Uveitis | 18670 | 9540 |
| Multiple valve disorder | 18084 | 10842 |
| Primary Malignancy: Liver | 17852 | 4702 |
| Immunodeficiencies | 17075 | 5889 |
| Hypertrophic Nasal Turbinates | 16713 | 14817 |
| Secondary Malignancy: Brain | 15639 | 6079 |
| Primary Malignancy: Bone | 15239 | 3893 |
| Pericardial Effusion | 14946 | 9584 |
| Secondary pulmonary hypertension | 14275 | 7193 |
| Primary Malignancy: Biliary Tract | 14152 | 3471 |
| Portal hypertension | 13793 | 8128 |
| Secondary Malignancy: Bone | 12958 | 4862 |
| Hepatic failure | 11139 | 5237 |
| Sick sinus syndrome | 10286 | 5627 |
| Secondary Malignancy: Lung | 8064 | 3608 |
| Angiodysplasia of colon | 7116 | 4165 |

| | | |
|---|---|---|
| Secondary Malignancy: Lymph Nodes | 6846 | 4075 |
| Primary Malignancy: Mesothelioma | 4267 | 994 |
| Enteropathic arthropathy | 3363 | 1125 |
| Tubulo-interstitial nephritis | 3052 | 1462 |
| Secondary Malignancy: Peritoneum | 1914 | 992 |
| Secondary Malignancy: Adrenal Gland | 274 | 171 |
| Secondary Malignancy: Bowel | 206 | 114 |
| Secondary Malignancy: Pleura | 157 | 54 |
| Primary Malignancy: Multiple Sites | 97 | 40 |

**Figure 2: Scree plot from MCA, for the first 30 dimensions**

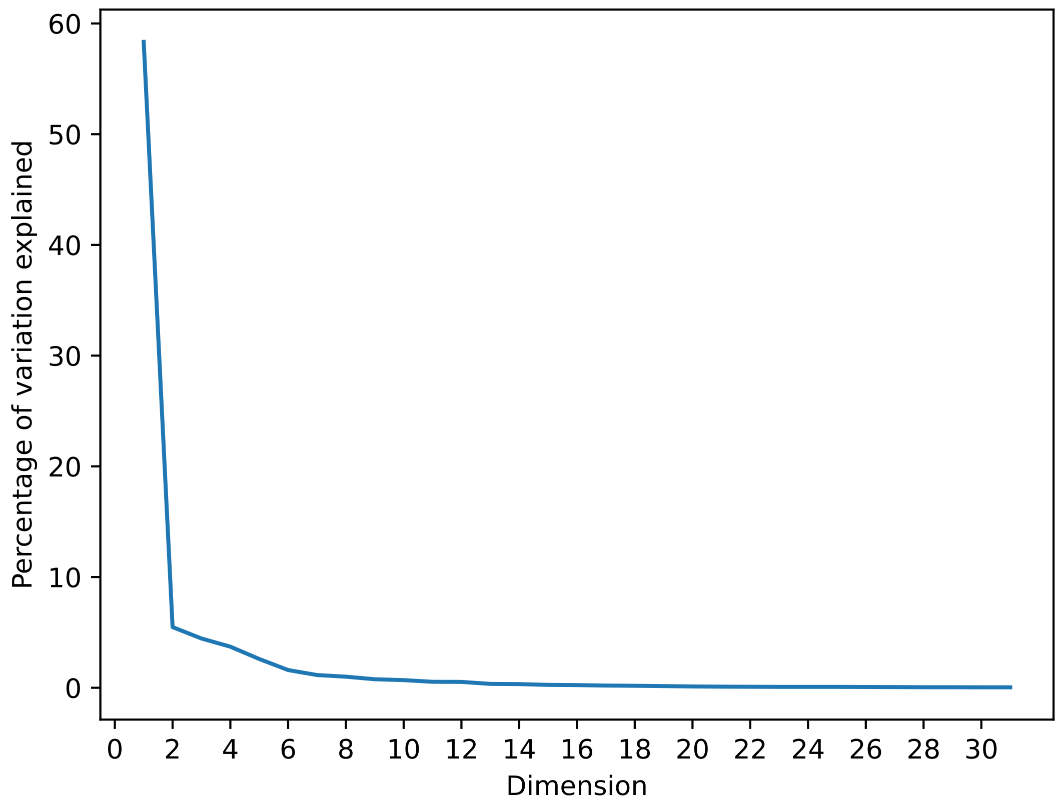

**Figure 3: Coordinates from the first two dimensions from MCA, coloured by disease system**

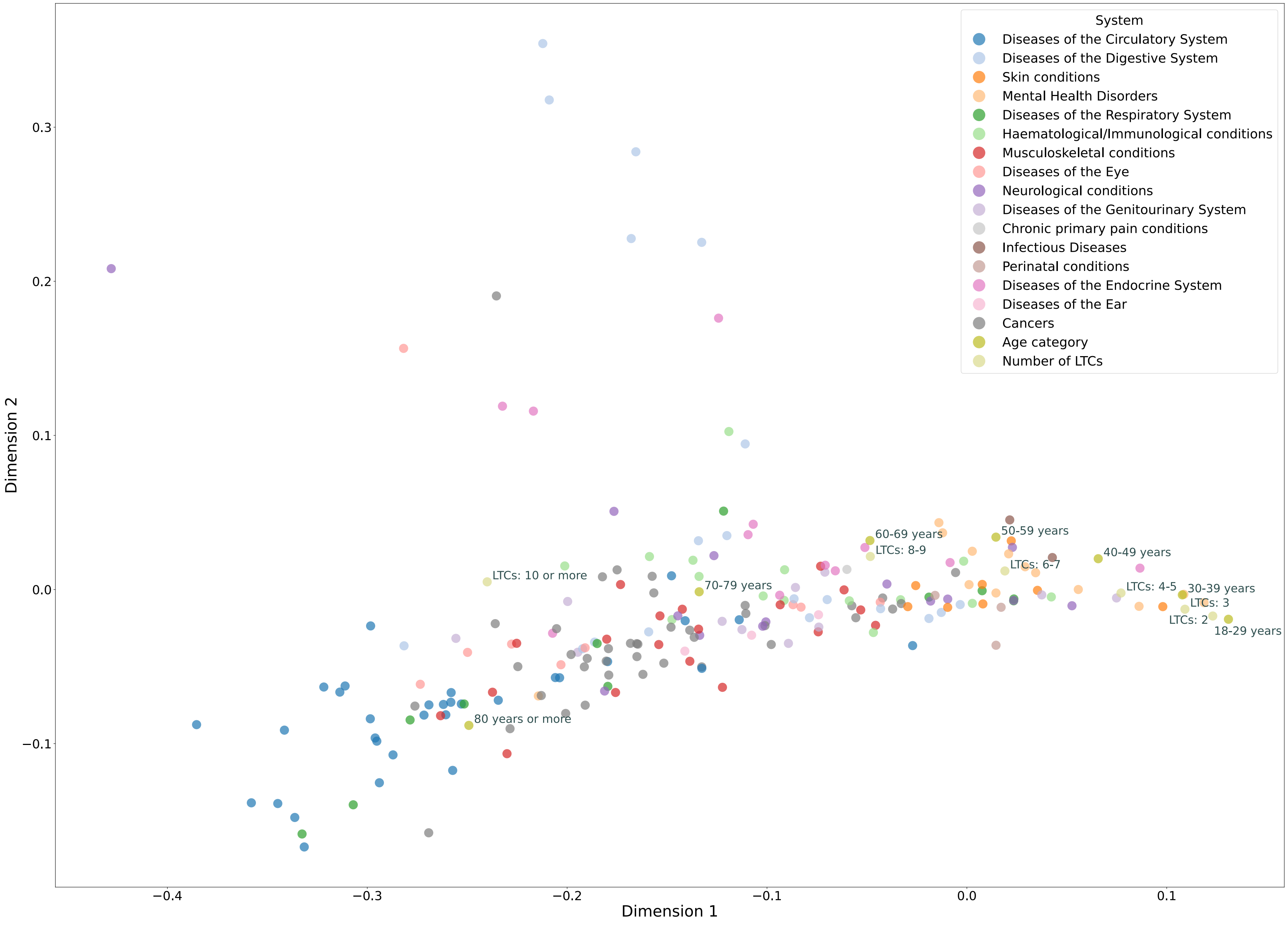


An interactive version of the figure is available at: https://tbeaney.github.io/MMclustering/mca_fig.html

**Embedding model hyperparameter selection**

We compared each of the GloVe, CBOW and SG models using the default parameter settings. We then altered each parameter in turn. For Word2vec models, we experimented with:

- Using vector sizes of 10 or 30. Vector dimensions ranging from 100-300 are commonly used, for vocabularies in the range of 10,000 or more. Given we were using only 212 unique diseases, suggested a vector size of 10 or 30 would be sufficient.
- Reducing the window size to 2 (from the default 5), given sequences were shorter than for many documents in natural language.
- Reducing the negative sampling windows to 2 (from the default 5) given shorter sequences.
- Reducing the down-sampling of frequent codes to 0 (from the default 0.001).

For GloVe models, we experimented with:

- Vector sizes of 10 or 30, as above.
- Reducing the window size to 2 (from the default 5), given sequences were shorter sequences.
- Learning rates of 0.01, 0.05 and 0.1.

In each case, we trialled models over 10 or 50 epochs for CBOW and SG, and steps for 10 to 150 for GloVe given that GloVe was computationally quicker to run. We ran CBOW and SG on unique (Table 3a) and multiple (Table 5a) codes and GloVe on unique (Table 4) and multiple (Table 6) codes. For the best performing CBOW and SG models, we then trialled runs over 10-50 epochs in steps of 10 to find the optimal number of epochs for unique (Table 3b) and multiple (Table 5b) codes. The best-performing models are highlighted in each table in bold.

**Table 3a: Performance of Word2vec models with different hyperparameters for unique disease codes only, ordered by performance**

| Model | Vector size | Window | Negative | Sampling | Epochs | Top 10 | Model | Vector size | Window | Negative | Sampling | Epochs | Top 10 |
|---|---|---|---|---|---|---|---|---|---|---|---|---|---|
| **CBOW** | **10** | **2** | **2** | **0.001** | **10** | **42.1** | **SG** | **10** | **2** | **2** | **0.001** | **10** | **47.6** |
| CBOW | 10 | 2 | 5 | 0.001 | 10 | 41.7 | SG | 10 | 5 | 2 | 0.001 | 10 | 46.2 |
| CBOW | 10 | 2 | 2 | 0.001 | 10 | 40.7 | SG | 10 | 2 | 2 | 0.001 | 10 | 46.0 |
| CBOW | 10 | 2 | 5 | 0.001 | 10 | 40.1 | SG | 10 | 5 | 2 | 0.001 | 10 | 46.0 |
| CBOW | 10 | 5 | 2 | 0.001 | 10 | 38.3 | SG | 10 | 2 | 5 | 0.001 | 10 | 44.9 |
| CBOW | 10 | 5 | 2 | 0.001 | 10 | 35.2 | SG | 10 | 5 | 5 | 0.001 | 10 | 44.3 |
| CBOW | 10 | 5 | 5 | 0.001 | 10 | 34.6 | SG | 10 | 5 | 5 | 0.001 | 10 | 44.1 |
| CBOW | 10 | 5 | 5 | 0.001 | 10 | 34.4 | SG | 10 | 2 | 5 | 0.001 | 10 | 44.1 |
| CBOW | 30 | 2 | 2 | 0 | 50 | 32.6 | SG | 30 | 2 | 2 | 0 | 50 | 33.0 |
| CBOW | 30 | 2 | 5 | 0.001 | 50 | 30.4 | SG | 30 | 2 | 2 | 0.001 | 50 | 32.6 |
| CBOW | 30 | 2 | 2 | 0 | 50 | 30.0 | SG | 30 | 5 | 2 | 0.001 | 50 | 32.4 |
| CBOW | 30 | 2 | 5 | 0 | 50 | 29.4 | SG | 30 | 2 | 5 | 0 | 50 | 32.2 |
| CBOW | 30 | 2 | 5 | 0.001 | 50 | 29.4 | SG | 30 | 2 | 2 | 0 | 50 | 31.8 |
| CBOW | 30 | 2 | 5 | 0 | 50 | 29.4 | SG | 30 | 5 | 2 | 0 | 50 | 31.6 |
| CBOW | 30 | 2 | 2 | 0.001 | 50 | 29.2 | SG | 30 | 5 | 2 | 0.001 | 50 | 31.6 |
| CBOW | 30 | 2 | 2 | 0.001 | 50 | 28.5 | SG | 30 | 2 | 5 | 0 | 50 | 31.0 |
| CBOW | 30 | 5 | 5 | 0.001 | 50 | 23.7 | SG | 10 | 2 | 5 | 0 | 10 | 30.8 |
| CBOW | 10 | 2 | 2 | 0 | 10 | 23.5 | SG | 30 | 5 | 2 | 0 | 50 | 30.6 |
| CBOW | 30 | 5 | 5 | 0 | 50 | 22.7 | SG | 30 | 2 | 2 | 0.001 | 50 | 30.2 |
| CBOW | 10 | 2 | 2 | 0 | 10 | 22.5 | SG | 10 | 2 | 5 | 0 | 10 | 30.0 |
| CBOW | 30 | 5 | 2 | 0.001 | 50 | 22.5 | SG | 10 | 5 | 5 | 0 | 10 | 30.0 |
| CBOW | 30 | 5 | 5 | 0.001 | 50 | 22.5 | SG | 30 | 5 | 5 | 0 | 50 | 30.0 |
| CBOW | 30 | 5 | 2 | 0 | 50 | 22.3 | SG | 10 | 2 | 2 | 0 | 10 | 29.8 |
| CBOW | 30 | 5 | 2 | 0 | 50 | 21.1 | SG | 30 | 2 | 5 | 0.001 | 50 | 28.9 |
| CBOW | 30 | 5 | 2 | 0.001 | 50 | 20.9 | SG | 30 | 5 | 5 | 0 | 50 | 28.9 |
| CBOW | 30 | 5 | 5 | 0 | 50 | 20.9 | SG | 10 | 2 | 2 | 0 | 10 | 28.7 |
| CBOW | 10 | 5 | 2 | 0 | 10 | 17.8 | SG | 10 | 5 | 5 | 0 | 10 | 28.1 |
| CBOW | 10 | 5 | 5 | 0 | 10 | 17.6 | SG | 30 | 2 | 5 | 0.001 | 50 | 27.9 |
| CBOW | 10 | 2 | 5 | 0 | 10 | 17.4 | SG | 10 | 5 | 2 | 0 | 10 | 27.3 |
| CBOW | 10 | 5 | 2 | 0 | 10 | 17.4 | SG | 30 | 5 | 5 | 0.001 | 50 | 27.1 |
| CBOW | 10 | 2 | 5 | 0 | 10 | 17.2 | SG | 10 | 5 | 2 | 0 | 10 | 26.9 |
| CBOW | 10 | 5 | 5 | 0 | 10 | 17.2 | SG | 30 | 5 | 5 | 0.001 | 50 | 25.5 |

Note: CBOW = continuous bag of words; SG = Skip-gram

**Table 3b: Run over different epochs for best performing models above**

| Model | Epochs | | | | |
|---|---|---|---|---|---|
| | 10 | 20 | 30 | 40 | 50 |
| SG | **47.6** | 47.0 | 46.2 | 46.4 | 46.0 |
| CBOW | **42.1** | 41.3 | 41.7 | 41.7 | 40.7 |

### Table 4: Performance of GloVe models with different hyperparameters for unique disease codes only, ordered by performance

| Vector size | Window | Learning rate | Epochs | Top 10 | Vector size | Window | Learning rate | Epochs | Top 10 |
|---|---|---|---|---|---|---|---|---|---|
| **30** | **2** | **0.1** | **100** | **51.4** | 30 | 5 | 0.05 | 30 | 32.2 |
| 30 | 2 | 0.1 | 150 | 50.6 | 30 | 2 | 0.01 | 100 | 31.4 |
| 30 | 2 | 0.1 | 50 | 50.6 | 30 | 5 | 0.01 | 150 | 31.2 |
| 30 | 5 | 0.1 | 50 | 50.2 | 10 | 2 | 0.05 | 100 | 31.0 |
| 30 | 2 | 0.1 | 30 | 49.2 | 30 | 5 | 0.05 | 20 | 30.0 |
| 30 | 5 | 0.1 | 150 | 48.6 | 30 | 2 | 0.05 | 10 | 28.9 |
| 30 | 5 | 0.1 | 100 | 48.4 | 10 | 5 | 0.05 | 50 | 28.5 |
| 30 | 2 | 0.1 | 20 | 47.8 | 30 | 5 | 0.05 | 10 | 26.9 |
| 30 | 5 | 0.1 | 30 | 47.8 | 30 | 5 | 0.01 | 100 | 25.3 |
| 30 | 2 | 0.05 | 150 | 46.6 | 10 | 5 | 0.05 | 20 | 25.1 |
| 30 | 2 | 0.05 | 100 | 46.4 | 10 | 2 | 0.05 | 50 | 24.7 |
| 30 | 5 | 0.1 | 20 | 45.8 | 10 | 5 | 0.05 | 30 | 24.5 |
| 30 | 2 | 0.1 | 10 | 45.5 | 10 | 2 | 0.05 | 30 | 24.3 |
| 30 | 5 | 0.05 | 150 | 44.7 | 30 | 2 | 0.01 | 50 | 23.5 |
| 10 | 2 | 0.1 | 100 | 43.9 | 10 | 2 | 0.05 | 20 | 23.3 |
| 10 | 2 | 0.1 | 150 | 43.9 | 10 | 2 | 0.05 | 10 | 22.7 |
| 10 | 5 | 0.1 | 100 | 43.7 | 10 | 5 | 0.05 | 10 | 17.6 |
| 10 | 5 | 0.1 | 150 | 43.7 | 30 | 2 | 0.01 | 30 | 16.6 |
| 30 | 5 | 0.05 | 100 | 42.1 | 30 | 5 | 0.01 | 50 | 15.8 |
| 30 | 5 | 0.1 | 10 | 41.5 | 10 | 2 | 0.01 | 150 | 14.4 |
| 10 | 2 | 0.1 | 50 | 41.3 | 30 | 2 | 0.01 | 20 | 13.6 |
| 10 | 5 | 0.1 | 50 | 40.7 | 30 | 5 | 0.01 | 30 | 12.8 |
| 30 | 2 | 0.05 | 50 | 40.5 | 30 | 2 | 0.01 | 10 | 12.1 |
| 10 | 5 | 0.1 | 30 | 37.9 | 10 | 2 | 0.01 | 100 | 11.7 |
| 10 | 2 | 0.1 | 30 | 37.7 | 30 | 5 | 0.01 | 20 | 11.1 |
| 30 | 2 | 0.05 | 30 | 37.4 | 10 | 5 | 0.01 | 150 | 10.9 |
| 10 | 2 | 0.1 | 20 | 37.0 | 30 | 5 | 0.01 | 10 | 9.9 |
| 30 | 5 | 0.05 | 50 | 37.0 | 10 | 5 | 0.01 | 100 | 7.9 |
| 10 | 5 | 0.1 | 20 | 36.0 | 10 | 2 | 0.01 | 50 | 7.5 |
| 10 | 2 | 0.05 | 150 | 35.4 | 10 | 2 | 0.01 | 30 | 6.5 |
| 10 | 5 | 0.05 | 150 | 35.2 | 10 | 5 | 0.01 | 50 | 6.5 |
| 30 | 2 | 0.05 | 20 | 33.4 | 10 | 5 | 0.01 | 30 | 6.3 |
| 10 | 2 | 0.1 | 10 | 33.2 | 10 | 2 | 0.01 | 10 | 6.1 |
| 10 | 5 | 0.1 | 10 | 33.0 | 10 | 5 | 0.01 | 20 | 6.1 |
| 10 | 5 | 0.05 | 100 | 32.8 | 10 | 2 | 0.01 | 20 | 5.9 |
| 30 | 2 | 0.01 | 150 | 32.4 | 10 | 5 | 0.01 | 10 | 5.1 |

### Table 5a: Performance of Word2vec models with different hyperparameters including recurrent codes, ordered by performance

| Model | Vector size | Window | Negative | Sampling | Epochs | Top 10 | Model | Vector size | Window | Negative | Sampling | Epochs | Top 10 |
|---|---|---|---|---|---|---|---|---|---|---|---|---|---|
| **CBOW** | **30** | **2** | **5** | **0** | **10** | **45.1** | **SG** | **30** | **5** | **2** | **0.001** | **50** | **55.3** |
| CBOW | 30 | 2 | 2 | 0.001 | 10 | 44.9 | SG | 30 | 2 | 5 | 0.001 | 50 | 55.3 |
| CBOW | 30 | 2 | 2 | 0 | 50 | 44.5 | SG | 30 | 2 | 2 | 0.001 | 10 | 55.3 |
| CBOW | 30 | 2 | 5 | 0 | 50 | 44.1 | SG | 30 | 2 | 5 | 0 | 10 | 54.3 |
| CBOW | 10 | 5 | 5 | 0.001 | 50 | 43.7 | SG | 30 | 5 | 2 | 0.001 | 10 | 54.3 |
| CBOW | 30 | 2 | 5 | 0.001 | 10 | 43.7 | SG | 30 | 2 | 2 | 0.001 | 50 | 54.3 |
| CBOW | 30 | 5 | 5 | 0 | 10 | 43.5 | SG | 30 | 2 | 2 | 0 | 10 | 53.6 |
| CBOW | 30 | 2 | 2 | 0 | 10 | 43.3 | SG | 30 | 2 | 5 | 0.001 | 10 | 52.8 |
| CBOW | 10 | 5 | 5 | 0.001 | 10 | 42.7 | SG | 30 | 2 | 2 | 0 | 50 | 52.8 |
| CBOW | 30 | 5 | 5 | 0 | 50 | 42.7 | SG | 30 | 5 | 5 | 0.001 | 10 | 52.0 |
| CBOW | 10 | 2 | 5 | 0.001 | 10 | 42.5 | SG | 30 | 5 | 2 | 0 | 10 | 51.8 |
| CBOW | 10 | 5 | 2 | 0.001 | 10 | 42.5 | SG | 30 | 5 | 2 | 0 | 50 | 51.6 |
| CBOW | 10 | 5 | 2 | 0.001 | 50 | 42.5 | SG | 30 | 5 | 5 | 0.001 | 50 | 51.6 |
| CBOW | 30 | 2 | 2 | 0.001 | 50 | 42.5 | SG | 30 | 2 | 5 | 0 | 50 | 51.4 |
| CBOW | 10 | 2 | 2 | 0.001 | 10 | 41.9 | SG | 30 | 5 | 5 | 0 | 50 | 48.6 |
| CBOW | 10 | 2 | 5 | 0 | 50 | 41.9 | SG | 30 | 5 | 5 | 0 | 10 | 48.4 |
| CBOW | 30 | 5 | 5 | 0.001 | 10 | 41.9 | SG | 10 | 2 | 2 | 0.001 | 10 | 47.2 |
| CBOW | 30 | 2 | 5 | 0.001 | 50 | 41.9 | SG | 10 | 5 | 2 | 0.001 | 10 | 47.2 |
| CBOW | 10 | 2 | 5 | 0.001 | 50 | 41.7 | SG | 10 | 5 | 5 | 0.001 | 10 | 47.2 |
| CBOW | 30 | 5 | 5 | 0.001 | 50 | 41.1 | SG | 10 | 2 | 5 | 0.001 | 10 | 46.6 |
| CBOW | 10 | 2 | 2 | 0.001 | 50 | 39.9 | SG | 10 | 2 | 5 | 0.001 | 50 | 46.0 |
| CBOW | 30 | 5 | 2 | 0.001 | 10 | 39.7 | SG | 10 | 2 | 2 | 0 | 50 | 45.5 |
| CBOW | 10 | 5 | 5 | 0 | 50 | 39.1 | SG | 10 | 2 | 2 | 0.001 | 50 | 45.3 |
| CBOW | 10 | 2 | 2 | 0 | 50 | 38.9 | SG | 10 | 5 | 2 | 0.001 | 50 | 45.3 |
| CBOW | 10 | 2 | 5 | 0 | 10 | 38.7 | SG | 10 | 5 | 5 | 0.001 | 50 | 44.9 |
| CBOW | 30 | 5 | 2 | 0.001 | 50 | 38.7 | SG | 10 | 2 | 2 | 0 | 10 | 44.7 |
| CBOW | 10 | 2 | 2 | 0 | 10 | 38.5 | SG | 10 | 2 | 5 | 0 | 10 | 44.3 |
| CBOW | 10 | 5 | 2 | 0 | 50 | 38.1 | SG | 10 | 2 | 5 | 0 | 50 | 43.9 |
| CBOW | 10 | 5 | 2 | 0 | 10 | 37.9 | SG | 10 | 5 | 2 | 0 | 10 | 43.5 |
| CBOW | 10 | 5 | 5 | 0 | 10 | 37.5 | SG | 10 | 5 | 2 | 0 | 50 | 43.1 |
| CBOW | 30 | 5 | 2 | 0 | 50 | 36.6 | SG | 10 | 5 | 5 | 0 | 10 | 42.3 |
| CBOW | 30 | 5 | 2 | 0 | 10 | 36.4 | SG | 10 | 5 | 5 | 0 | 50 | 41.1 |

Note: CBOW = continuous bag of words; SG = Skip-gram

### Table 5b: Run over different epochs for best performing models above

| Model | Epochs | | | | |
|---|---|---|---|---|---|
| | 10 | 20 | 30 | 40 | 50 |
| SG | 54.3 | **56.1** | 55.5 | 55.3 | 55.3 |
| CBOW | **45.1** | 42.5 | 44.7 | 43.1 | 44.1 |

**Table 6: Performance of GloVe models with different hyperparameters including recurrent codes, ordered by performance**

| Vector size | Window | Learning rate | Epochs | Top 10 | Vector size | Window | Learning rate | Epochs | Top 10 |
|---|---|---|---|---|---|---|---|---|---|
| **30** | **5** | **0.1** | **100** | **51.4** | 30 | 2 | 0.01 | 150 | 33.4 |
| 30 | 2 | 0.1 | 150 | 51.4 | 10 | 5 | 0.1 | 10 | 33.0 |
| 30 | 5 | 0.1 | 150 | 51.2 | 30 | 5 | 0.05 | 20 | 32.8 |
| 30 | 5 | 0.1 | 200 | 51.0 | 10 | 5 | 0.05 | 100 | 32.4 |
| 30 | 2 | 0.1 | 100 | 50.8 | 30 | 2 | 0.05 | 20 | 31.6 |
| 30 | 2 | 0.1 | 200 | 50.6 | 30 | 5 | 0.01 | 200 | 31.2 |
| 30 | 2 | 0.1 | 50 | 50.2 | 30 | 2 | 0.01 | 100 | 31.0 |
| 30 | 5 | 0.1 | 50 | 50.2 | 30 | 5 | 0.01 | 150 | 30.6 |
| 30 | 2 | 0.1 | 30 | 49.6 | 30 | 2 | 0.01 | 200 | 29.8 |
| 30 | 5 | 0.1 | 30 | 49.4 | 10 | 2 | 0.05 | 50 | 29.6 |
| 30 | 2 | 0.1 | 20 | 49.0 | 30 | 5 | 0.05 | 10 | 26.5 |
| 30 | 2 | 0.05 | 200 | 48.6 | 30 | 5 | 0.01 | 100 | 26.1 |
| 30 | 5 | 0.05 | 200 | 48.2 | 10 | 2 | 0.05 | 30 | 25.3 |
| 30 | 5 | 0.05 | 150 | 47.6 | 10 | 5 | 0.05 | 50 | 24.7 |
| 30 | 5 | 0.1 | 20 | 47.6 | 10 | 2 | 0.05 | 20 | 23.3 |
| 30 | 2 | 0.05 | 150 | 47.4 | 10 | 5 | 0.05 | 20 | 22.9 |
| 10 | 2 | 0.1 | 200 | 46.2 | 10 | 5 | 0.05 | 30 | 22.5 |
| 30 | 2 | 0.1 | 10 | 46.2 | 30 | 2 | 0.01 | 50 | 21.7 |
| 10 | 2 | 0.1 | 150 | 46.0 | 30 | 2 | 0.05 | 10 | 21.3 |
| 30 | 2 | 0.05 | 100 | 45.7 | 10 | 2 | 0.05 | 10 | 20.2 |
| 10 | 2 | 0.1 | 100 | 45.5 | 10 | 5 | 0.05 | 10 | 20.0 |
| 10 | 5 | 0.1 | 150 | 45.5 | 30 | 5 | 0.01 | 50 | 18.8 |
| 30 | 5 | 0.1 | 10 | 45.3 | 10 | 2 | 0.01 | 200 | 15.8 |
| 10 | 5 | 0.1 | 200 | 45.1 | 30 | 2 | 0.01 | 30 | 15.0 |
| 30 | 5 | 0.05 | 100 | 44.9 | 30 | 5 | 0.01 | 30 | 14.8 |
| 10 | 5 | 0.1 | 100 | 44.7 | 10 | 5 | 0.01 | 200 | 13.8 |
| 10 | 2 | 0.1 | 50 | 43.3 | 10 | 2 | 0.01 | 150 | 13.4 |
| 30 | 2 | 0.05 | 50 | 42.5 | 30 | 2 | 0.01 | 20 | 12.5 |
| 30 | 5 | 0.05 | 50 | 41.9 | 30 | 5 | 0.01 | 20 | 12.5 |
| 10 | 2 | 0.1 | 30 | 41.7 | 10 | 2 | 0.01 | 100 | 11.7 |
| 10 | 5 | 0.1 | 50 | 41.5 | 10 | 2 | 0.01 | 50 | 11.3 |
| 10 | 2 | 0.05 | 200 | 40.3 | 10 | 5 | 0.01 | 150 | 10.7 |
| 10 | 2 | 0.1 | 20 | 39.9 | 30 | 2 | 0.01 | 10 | 10.5 |
| 30 | 5 | 0.05 | 30 | 38.9 | 30 | 5 | 0.01 | 10 | 10.3 |
| 10 | 5 | 0.1 | 30 | 38.7 | 10 | 2 | 0.01 | 30 | 9.5 |
| 10 | 5 | 0.05 | 200 | 38.3 | 10 | 2 | 0.01 | 20 | 8.9 |
| 10 | 2 | 0.05 | 150 | 38.1 | 10 | 2 | 0.01 | 10 | 8.5 |
| 30 | 2 | 0.05 | 30 | 38.1 | 10 | 5 | 0.01 | 100 | 7.9 |
| 10 | 2 | 0.05 | 100 | 36.4 | 10 | 5 | 0.01 | 50 | 7.1 |
| 10 | 5 | 0.1 | 20 | 35.6 | 10 | 5 | 0.01 | 20 | 6.5 |
| 10 | 5 | 0.05 | 150 | 35.2 | 10 | 5 | 0.01 | 30 | 6.5 |
| 10 | 2 | 0.1 | 10 | 35.0 | 10 | 5 | 0.01 | 10 | 5.9 |

**Figure 4: Comparison performance of each embedding model using different number of nearest neighbours of 2, 5, 10 and 20**

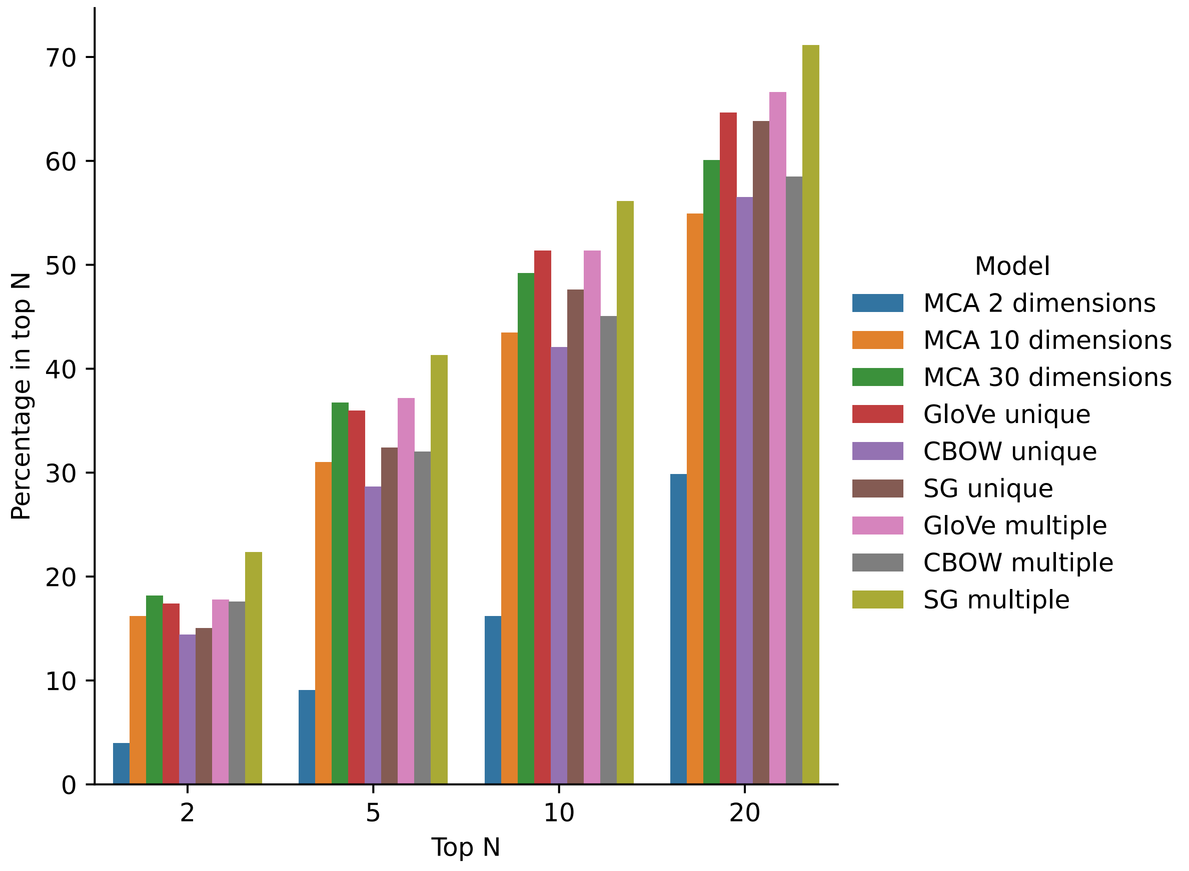

**Figure 5: Mean Jaccard similarity of the set of two diseases for each patient assigned to a disease cluster, using MCA-30 embeddings (panel A) and SG-M embeddings (panel B)**

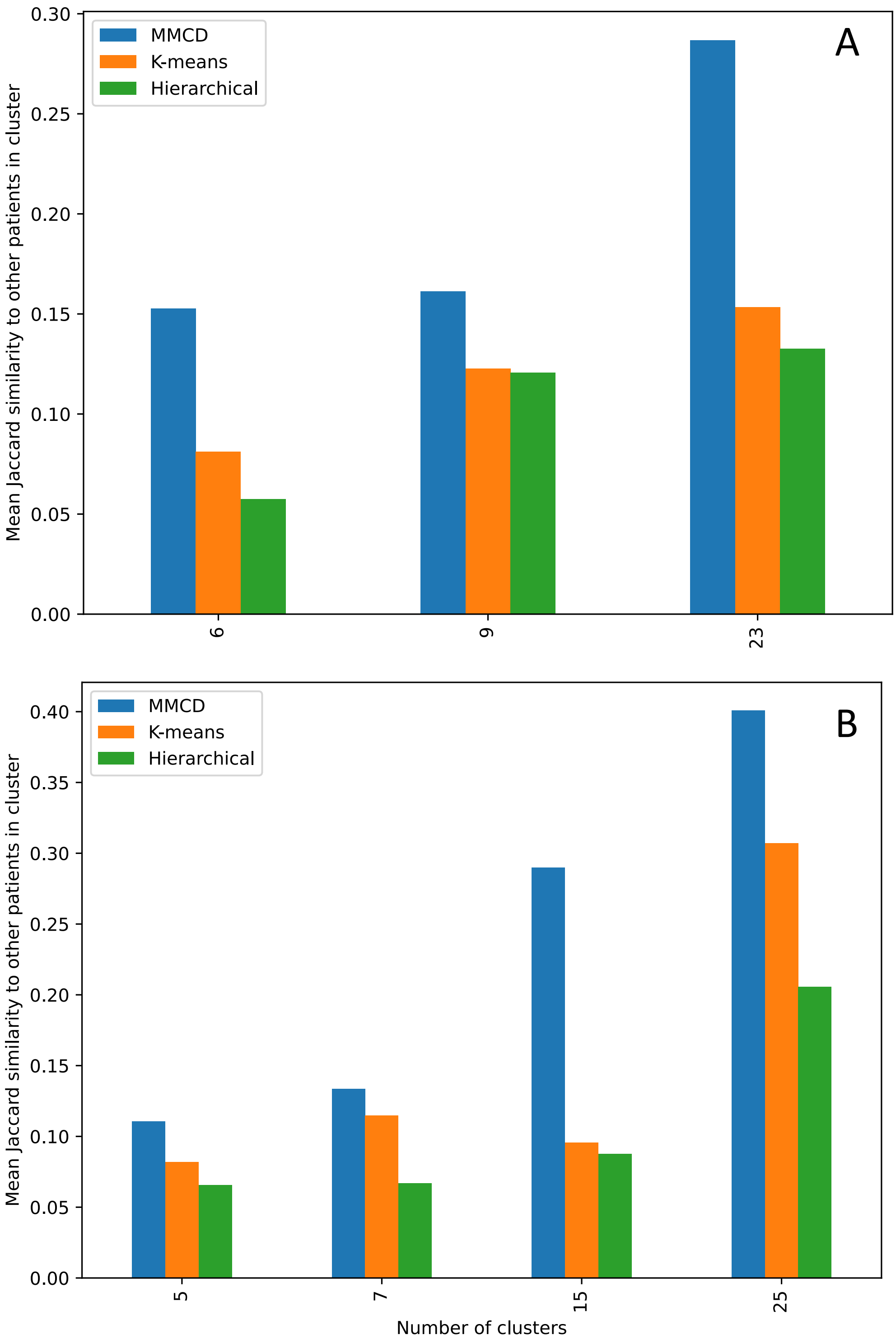

**Table 7A: Normalised variation of information (NVI) comparing assignment of diseases to clusters from MCA-30 to chapters in ICD-10**

| Number of clusters | NVI |
|---|---|
| 23 | 0.60 |
| 9 | 0.70 |
| 6 | 0.75 |

**Table 7B: Normalised variation of information (NVI) comparing assignment of diseases to clusters from SG-M to chapters in ICD-10**

| Number of clusters | NVI |
|---|---|
| 25 | 0.55 |
| 15 | 0.62 |
| 7 | 0.68 |
| 5 | 0.75 |

**Table 8: Ratio of observed to expected prevalence of co-occurrence for each pair of conditions in the thyroid and other malignancy cluster (SG-M at a 15-cluster resolution)**

| Disease | Primary Malignancy: Brain | Primary Malignancy: Melanoma | Primary Malignancy: Testis | Primary Malignancy: Thyroid |
|---|---|---|---|---|
| Primary Malignancy: Melanoma | 2.29 | | | |
| Primary Malignancy: Testis | 1.96 | 1.17 | | |
| Primary Malignancy: Thyroid | 1.61 | 2.35 | 0.89 | |
| Thyroid Disease | 1.61 | 1.36 | 0.45 | 9.29 |
